# Carriers of *SCN9A* variants linked to inherited and acquired pain syndromes show no alteration in the prevalence of pain or analgesic usage in the UK Biobank cohort

**DOI:** 10.1101/2025.03.12.25323817

**Authors:** Graeme W.T. Newton, Corin Charman, Aidan P. Nickerson, Keith G. Phillips, Achim Kless, James P Dunham, Anthony E Pickering

## Abstract

The voltage-gated sodium channel NaV1.7, encoded by the *SCN9A* gene, is integral to nociceptor excitability and pain sensation. Multiple gain-of-function *SCN9A* variants have been reported to cause autosomal dominant painful channelopathies, including primary erythromelalgia and paroxysmal extreme pain disorder, and they have been linked to the pathogenesis of painful small fibre neuropathies. The prevalence and impact on carriers of these gain-of-function NaV1.7 variants in the wider population has yet to be explored. A literature search was performed to identify pathogenic *SCN9A* variants linked to painful channelopathies. *SCN9A* variants that were associated with small fibre neuropathy were also included if they had *in vitro* electrophysiological evidence of gain-of-function. We investigated the prevalence of these *SCN9A* variants in the UK Biobank cohort (using the 470K whole-exome sequencing dataset). Logistic regression was used to examine whether carrying pathogenic variants was associated with chronic pain, neuropathic pain, and analgesic or anti-neuropathic drug prescription (adjusting for age, sex, and the first 10 genetic principal components).

A total of 59 putative pathogenic gain-of-function variants in *SCN9A* were identified from the literature review, of which 20 were found in the UK Biobank cohort (in over 148,000 carriers, >29%). Logistic regression analysis was feasible for 11 of the 20 variants which each had ≥50 heterozygous carriers and were not in linkage disequilibrium with another putative pathogenic mutation. None of these NaV1.7 mutation carrier groups had evidence of an increased risk of chronic or neuropathic pain. Similarly, no evidence of an increase in prescriptions for opioid analgesics or anti-neuropathic pain medications was found. Subsequent pathogenicity prediction analysis using AlphaMissense, CADD, and EVE indicated that *SCN9A* variants called in UK Biobank (20/59) were significantly less likely to be rated pathogenic than those that were absent from the cohort (39/59).

These findings show that almost one third of the previously reported pathogenic *SCN9A* gain-of-function variants are likely to be benign from a pain perspective. This study challenges the previously assumed high penetrance of these *SCN9A* gain-of-function variants for chronic pain and we make recommendations to optimise the evidence-based criteria for genetic association studies investigating the role of ion channel variants in the pathogenesis of pain.

## Introduction

The NaV1.7 voltage-gated sodium channel plays a crucial role in determining neuronal excitability in the peripheral nervous system. The alpha subunit of the NaV1.7 voltage gated sodium channel is encoded by the *SCN9A* gene located on chromosome 2. The alpha subunit has 4 domains each consisting of 6 transmembrane helical segments with a pore-forming loop^1^. The fourth transmembrane segment (S4) in each domain acts as a voltage sensor. The fifth and sixth transmembrane segments (S5&6) form the channel pore. The link between the third and fourth domains forms the fast inactivation gate, which blocks the channel pore after depolarisation, preventing further sodium influx^1^.

The influx of Na^+^ through voltage-gated sodium channels is a crucial determinant of neuronal excitability through the generation of action potentials^2^. NaV1.7 is considered to function as a threshold channel in primary afferent nociceptors, meaning it is crucial for the initiation of action potentials in response to generator potentials^3^. NaV1.7 has been causally linked to multiple human pain-related channelopathies including both congenital insensitivity to pain and severe chronic pain conditions like erythromelalgia and paroxysmal extreme pain disorder^3^ supporting its critical role in nociception. Congenital insensitivity to pain is extremely rare and some of these patients have been shown to have homozygous loss of function variants in NaV1.7 ^4^ leading to a failure of nociceptors to generate action potentials. These patients sustain repeated debilitating injuries due to the loss of pain sensation including long bone fractures, Charcot joints, corneal and oral ulceration. As a consequence of these recurrent injuries, and associated infections, these patients have a greatly shortened life expectancy.

In contrast, gain-of-function NaV1.7 mutations have been associated with rare inherited chronic pain conditions such as erythromelalgia, where carriage of a single copy of the variant being sufficient to cause the disease (heterozygous). This was first described by Silas Weir Mitchell who published a collection of 15 case studies of patients experiencing redness and severe pain in the extremities^5^. His observations of pain episodes being aggravated by physical exercise or warmth, their temporary relief with cooling, their variable severity, and refractoriness to attempts at treatment still apply today. Patients with primary erythromelalgia typically present in early childhood and suffer from extreme pain that is often not helped by either analgesics or by anti-neuropathic pain medications^6,7^. It can be inherited in an autosomal dominant pattern or can occur *de novo* with new NaV1.7 mutations in individuals without a family history^8^. Many unique *SCN9A* putative gain-of-function variants have been causally linked to erythromelalgia in the published literature^3^. There appears to be an association between disease presentation and the biophysical properties of the mutant NaV1.7 channels with more severe electrophysiological changes linked to earlier disease onset^9,10^. In a cohort study of 13 erythromelalgia patients, some of the pain attacks were triggered by exercise, heat, routine household tasks, and contact with clothing but the majority of attacks occurred without any obvious trigger^11^.

Paroxysmal Extreme Pain Disorder was first reported under the term “familial rectal pain” ^12^as the index infant presented with episodic rectal, ocular, and mandibular pain and redness in the affected areas with a family history of such symptoms. The clinical presentation is highly variable, even within the same family, and patients do not always present with rectal symptoms. As such, the condition was later renamed paroxysmal extreme pain disorder (PEPD) to better reflect the variability in clinical presentation^13^. PEPD is comparatively rarer than erythromelalgia and is consequently less well phenotyped^14^. Gene-linkage studies initially tied PEPD to chromosome 2 and given the similarities in episodic burning pain and redness, the “erythromelalgia gene”, *SCN9A*, was sequenced^13^. This identified gain-of-function NaV1.7 mutations in these PEPD patients and again identified both autosomal dominant inheritance patterns and *de novo* mutations.

Small fibre neuropathy (SFN) is typically an acquired disease. Diabetes, exposure to toxins, and immunological disorders are the most common aetiologies, although many cases are idiopathic. There is increasing evidence for a genetic component in both idiopathic SFN and SFN secondary to other causes^15^. Recent studies have reported associations with a number of *SCN9A* variants which are found at numerically increased rates in cohorts of small-fibre neuropathy patients^16,17^. Several of these variants have been shown to exhibit gain of function electrophysiological properties, supporting the hypothesis that they may be contributing to SFN pathogenesis and potentially to pain.

Functional studies of NaV1.7 mutations have been performed on many of the *SCN9A* variants identified in patients with erythromelalgia, PEPD, and small fibre neuropathy. These are typically expression studies in heterologous cell lines, often using patch clamp recording to examine the channel biophysics. Typically, NaV1.7 mutations associated with erythromelalgia are characterised by a hyperpolarising shift in activation, effectively decreasing the action potential threshold to a level nearer the neuron’s resting membrane potential. In contrast, NaV1.7 mutations linked to PEPD frequently result in a depolarising shift in fast inactivation, requiring a more positive membrane potential for channel inactivation^18^. One particularly extreme phenotype of a PEPD mutation (L245V) causes such a large depolarising shift in inactivation that there is a persistent Na^+^ conductance, potentially leading to spontaneous firing^19,20^. Functional studies of human nociceptors *in vivo* are challenging, but some erythromelalgia patients have been studied with microneurography. Spontaneous activity and reduced activity-dependent slowing of C-fibres were identified in an erythromelalgia patient carrying the I228M mutation^21^. An erythromelalgia patient with NaV1.7 I848T mutation was also studied ^21^ which showed the patient’s C-fibres exhibited aberrant responses to electrical stimulation suggested to reflect a large axonal depolarisation^21^.

This cumulative weight of genetic and functional evidence that NaV1.7 is linked to inherited pain conditions is well accepted by the field^3,18,22^. This has led to efforts to generate analgesic therapies targeting this channel with 3 different NaV1.7 small molecule inhibitors having completed phase I and II clinical trials (PF-05089771 (Pfizer), AZD3161 (AstraZeneca), Vixotrigine (Biogen)). Although NaV1.7 has been a challenging target due to the conservation of structure with other voltage-gated sodium channels ^23,24^ there are still active industry programmes developing small molecule therapeutics and also using RNA manipulation methods to alter the expression of the protein^25^.

The studies of NaV1.7 to date have focussed mostly on these putative pathological variants in the setting of rare inherited conditions or small fibre neuropathy. The prevalence of these mutations in the wider general population and the potential association with pain phenotypes has not been systematically examined as the identification of these rare variants in the population cohorts was until recently not feasible – a situation that has now changed with the increasing availability of whole exome or whole genome sequencing data. In this study, the UK Biobank 470k whole exome sequencing dataset is explored to identify participants carrying any of the published erythromelalgia or paroxysmal extreme pain disorder mutations, as well as confirmed gain-of-function NaV1.7 mutations identified in patients with small fibre neuropathy. The study aimed to test the hypothesis that these gain-of-function, putatively pathogenic, NaV1.7 variants are associated with chronic pain by evaluating their relationship to pain phenotypes, analgesic and anti-neuropathic pain medication prescriptions, degree of evolutionary conservation, and pathogenicity predictions from clinical genetics databases and from machine learning models.

The voltage-gated sodium channels Nav1.8 and 1.9 are similarly thought to be implicated in nociception, although less evidence exists to support these links.Nav1.8 is encoded by the SCN10A gene, and is selectively expressed in the DRG and trigeminal ganglion neurons, where it tends to localise at free nerve endings. The channel has been shown to be involved in nociception and thus, may also be implicated in painful neuropathies; however, there exists substantially less evidence than for Nav1.7. Some evidence has suggested that gain-of-function mutations in SCN10A may be linked to small-fibre neuropathy, a disease causing burning or shooting pain in the extremities. Loss-of-function SCN10A mutations are similarly poorly understood. Two separate papers have identified loss-of-function variants of SCN10A that result in completely non-functional proteins, yet are associated with small-fibre neuropathy in three patients. The mechanism of this remains unclear.

Nav1.9 is encoded by SCN11A, identified as functionally expressed in DRG neurons, the trigeminal ganglion neurons, and intrinsic myenteric neurons. In primary sensory neurons, the channel tends to localise in the somata and free nerve endings. This is thought to be implicated in nociception, and the channel has been linked to painful channelopathies, however, like Nav1.8, the evidence is relatively limited. Most variants isolated have been gain-of-function, and associated with two painful channelopathies – familial episodic pain syndromes (FEPS) and small fibre neuropathy. FEPS comprises four subtypes, and is characterised by childhood onset of extreme pain episodes, normally in the arms and legs, the severity of which often attenuating with age (36,37). Conversely, no loss-of-function SCN11A variants have been identified.

## Materials and methods

### Identification of SCN9A Pathogenic Variants

Mutations linked to painful channelopathies in the NaV1.7 protein were identified from UniProt^26^. PubMed was searched for the primary research articles associated with these mutations using the term “SCN9A OR NaV1.7 AND *protein mutation name*”, for example, “SCN9A OR NaV1.7 AND I228M”. Additional follow-up searches in PubMed were conducted to ensure that any mutations missing from UniProt were captured. Reviews of NaV1.7 channelopathy mutations were also consulted as part of this process ^3,27-30^ with snowball searches of the cited literature.

Mutations reported in the primary literature as pathogenic for erythromelalgia and PEPD were included, both with and without electrophysiological gain-of-function evidence. Mutations reported as being associated with idiopathic small-fibre neuropathy were included only if there was gain-of-function electrophysiological evidence for the mutation. This was because several small fibre neuropathy cohort studies have reported an increased prevalence of many *SCN9A* variants but without formal statistical testing^31,32^. A mutation was considered to have been shown to confer gain-of-function (compared to control in any transfected cell culture model) if there was a statistically significant change in any of the following parameters from whole-cell patch clamp: activation threshold, slow inactivation, fast inactivation, or resting membrane potential.

Two additional NaV1.7 mutations, N641Y and K655R, were reported to be potentially pathogenic for epilepsy and had gain-of-function electrophysiological evidence^33^. As such, these mutations were included as a potential positive control to see whether this epileptogenic gain-of-function translated into a pain phenotype, or whether any associations would be identified with prescription sodium channel blockers, including many antiepileptic drugs.

### Variant Pathogenicity Predictions

To complement the functional phenotypes derived from UK Biobank, we also obtained variant pathogenicity predictions from ClinVar, UniProt, CADD, EVE, and AlphaMissense. ClinVar is a public archive of user-submitted associations between genetic variation and phenotypes, such as reports of results from patients referred for genetic testing^34^. UniProt is a public archive of protein sequence and annotation data from manually and computer-curated datasets^26^. Combined Annotation Dependent Depletion (CADD) scores are produced by a machine learning model that includes over 60 input parameters spanning evolutionary conservation to tissue expression^35^. The scores are then ranked and scaled between 0 – 99, with higher scores meaning a variant is likely more deleterious. The scaling is logarithmic, a score >10 meaning the mutation is in the top 10%, and scores >20 corresponding to being in the top 1%. The authors suggest a threshold of 15 for identifying potentially pathogenic variants (top 5%). The Evolutionary model of Variant Effect (EVE) score is generated by a machine learning model that predicts pathogenicity based on over 140k species amino acid sequence data – it scores between 0 and 1, and the predicted pathogenicity threshold is 0.7^36^. AlphaMissense is a machine-learning model that predicts the pathogenicity of missense mutations based on the conservation of protein sequence, rarity, and structural context. Like EVE, it scores between 0 and 1, and the predicted pathogenicity threshold is 0.7^37^.

### Protein Sequence Alignment and Identification of Paralogs

Conserved amino acids in an ion channel protein sequence are more likely to be essential for function and therefore mutations affecting these sites are more likely to be deleterious^38^. Multiple sequence alignment was performed to assess the degree of conservation for individual amino acid residues of NaV1.7. Protein sequences of NaV1.7 from human (*Homo sapiens*), chimpanzee (*Pan troglodytes*), rhesus macaque (*Macaca mulatta*), cow (*Bos taurus*), pig (*Sus scrofa*), dog (*Canis lupus familiaris*), rabbit (*Oryctolagus cuniculus*), guinea pig (*Cavia porcellus*), rat (*Rattus norvegicus*), and mouse (*Mus musculus*) were downloaded from NCBI RefSeq for interspecies sequence alignment. Protein sequences of the NaV1.1 – NaV1.9 and NaX channels were downloaded from NCBI RefSeq for intraspecies sequence alignment. Multiple protein sequence alignment was performed using the MUSCLE algorithm with default settings implemented in Jalview^39,40^. Sequence alignment scores are presented as integers from 0 – 10, representing the number of species or NaV channels where this amino acid was conserved.

Another method to triangulate the predictions of pathogenicity is through the comparative analysis of paralogues of each variant in the other human NaVs where there is strong sequence conservation in the functionally important domains^41^. Each NaV1.7 pathogenic variant was explored for paralogs that had been linked to other diseases, such as epilepsy or cardiac conduction deficits, and whether these had been shown to cause gain of function (https://scn-viewer.broadinstitute.org/).

### UK Biobank study

The UK Biobank cohort contains 502,394 participants; 273,316 (54.4%) are female, and 229,078 (45.6%) are male. The study enrolled participants aged 37 – 73 from 2006 to 2010^42^. This age range was selected as participants would be at increased risk of developing diseases such as cancer, heart disease, diabetes, and dementia as they aged over the following decades. Accordingly, the age range at the time of this study is between 52 and 89. Participants have undergone genetic sequencing, detailed phenotypic screening, and had their NHS hospital inpatient and prescription data linked. In 2022, the final whole exome sequencing dataset for the UK Biobank cohort was released^43^.

### Sequencing Data Processing and Variant Annotation

The UK Biobank Research Analysis Platform was used to extract whole exome sequencing data. The *SCN9A* gene region was extracted using PLINK 2 (www.cog-genomics.org/plink/2.0/) and then used to calculate the number of heterozygous and homozygous minor allele calls and convert the sequencing data to the vcf format^44^. Ensembl Variant Effect Predictor release 113.0 was used to annotate VCF files^45^.

### Pain Phenotype Categorisation

The UK Biobank Research Analysis Platform was also used to extract the phenotype data to give broad coverage of different types of chronic pain and from data gathered at different time points. A total of 7 binary phenotypes were generated using a combination of data from the original assessment at the cohort’s inception, the experience of pain questionnaire (EoPQ,^46^), and prescription history from the electronic health record linkage. In the original recruitment assessment, participants were considered positive for the “chronic pain” phenotype if they had pain for more than 3 months in any specific body area or more generally (**Supplementary Table 1**). A participant was considered positive for the “neuropathic pain” phenotype if they scored 3 or greater, in the 7 questions derived from the DN4 ^47^ in the EoPQ which was completed in 2019 (**Supplementary Table 2**). It has already been used to explore the epidemiology of neuropathic pain in the UK Biobank cohort^46^. The DN4 was chosen as it may be more specific in detecting neuropathic pain that would be predicted for the small-fibre neuropathy and erythromelalgia/PEPD NaV1.7 mutations.

To extract analgesic and anti-neuropathic pain prescription phenotypes, 56 million prescription events between 1986 - 2018 from 222,065 participants were queried using PySpark in an Apache Spark cluster on the UK Biobank Research Analysis Platform. Regular expression patterns were used to extract all possible entries for a particular drug, regardless of dose or prescription frequency (**Supplementary Table 3**). A participant was considered positive for a prescription drug phenotype if they had ever been prescribed a drug within that group. For example, if a participant had been prescribed carbamazepine, they were considered positive for the *“NaV_blocker”* phenotype. The prevalence of patients having ever had prescriptions for NaV channel blockers was *3.0%* (*4,929* participants); gabapentinoids *7.3%* (*16,132* participants); tricyclic antidepressants (TCAs) was *18.9%* (*42,012* participants); strong opioids was *4.0%* (*8,881* participants) and dual-mechanism opioids was *13.1%* (*29,108* participants).

### Statistical Analysis

The primary objective of the statistical analysis was to investigate the association between specific *SCN9A* gain-of-function mutations and the likelihood of experiencing chronic pain, neuropathic pain, or receiving analgesic and anti-neuropathic drug prescriptions. Binomial logistic regression models were used to examine the relationships between the outcome and predictor variables while adjusting for age, sex and the first 10 genetic principal components^48^.

Each participant was assigned to a single NaV1.7 mutation group. Participants without any of the 59 potentially pathogenic gain-of-function NaV1.7 mutations identified through the literature search were designated the control population (462,158 people). Participants who were heterozygous or homozygous for one pathogenic NaV1.7 mutation were assigned to the corresponding mutation group.

Participants that carried multiple pathogenic NaV1.7 mutations were removed from the analysis (*N* = 3,101). The majority of this was driven by the so called *“Neanderthal” SCN9A* haplotype (includes the M932L and V991L mutations), which has been linked to slightly increased pain sensitivity^49,50^. Individual mutation NaV1.7 groups with fewer than 50 carriers were also removed from the analysis as power calculations indicated that smaller groups would be insufficiently powered to reliably detect an OR of 1.5. This moderate OR would correspond to an increase in prevalence of around 10% from the observed control chronic pain rate of 41.7% to 51.8%. For a Fisher’s exact test, assuming an alpha 0.05, control population of 462,158, and a mutation group size of 50, the β (power) was 32.9% based on 10,000 simulations. As a sensitivity analysis the individual groups with fewer than 50 carriers were aggregated and analysed.

Participants were removed at the model level if data were missing for the outcome variable. The models were checked for multicollinearity. All logistic regression models were generated using *“glm()”* function from the base R package *“stats”* R version 4.3.2 (R Core Team 2023) and followed the same form, where *p* represented the probability of experiencing the phenotype being tested, and β represents model coefficients.

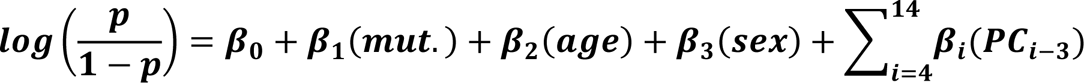

*P* values were adjusted at the model level to correct for multiple testing (7 logistic regressions) using the False Discovery Rate (FDR) method as implemented in the *“p.adjust()”* function. The alpha level (*P* value threshold) was set at 0.05. An odds ratio (OR) of > 1 indicated an increased likelihood of experiencing the phenotype. ORs are presented with the associated 95% confidence intervals.

The results from logistic regression of each putative pathogenic NaV1.7 mutation with ≥ 50 carriers in the UK Biobank are presented with additional context from the primary literature on three of the mutations with the greatest weight of prior evidence implicating them as associated with an inherited pain condition. Raw outputs from logistic regression models are presented in **Supplementary Table 4**.

## Data availability

The outputs of the analysis are provided in the paper and supplementary materials. Researchers can access the data from the UK Biobank cohort by applying on https://www.ukbiobank.ac.uk/enable-your-research/apply-for-access.

The code to reproduce this analysis is freely available at https://github.com/graemenewton/Newton_2025_NaV_UKB.

## Results

A total of 59 putative pathogenic *SCN9A* variants were identified from the literature search (**Supplementary Table 5**). Almost all of the reported erythromelalgia causal variants (*33 / 35,* 94%) and the large majority of the reported PEPD causal variants (*9 / 12, 75%*) had gain-of-function confirmed using electrophysiology (**Supplementary Table 5**). Similarly, all ten included SFN variants had undergone functional studies and were found to produce a gain-of-function (**Supplementary Table 5**). These putative pathogenic mutations, caused by amino acid substitutions, are dispersed widely throughout the NaV1.7 protein (**Figure 1A**). Every domain (D1-4) of NaV1.7 contains pathogenic mutations, as well as all segments (S1-6) and the pore-forming loop (contained within the S5-S6 linker). Calculating the rate of mutations per amino acid in each domain reveals that the S3-S4 linker, S4-S5 linker, and S5 are relatively enriched (**Figure 1B**).

**Figure 1.**
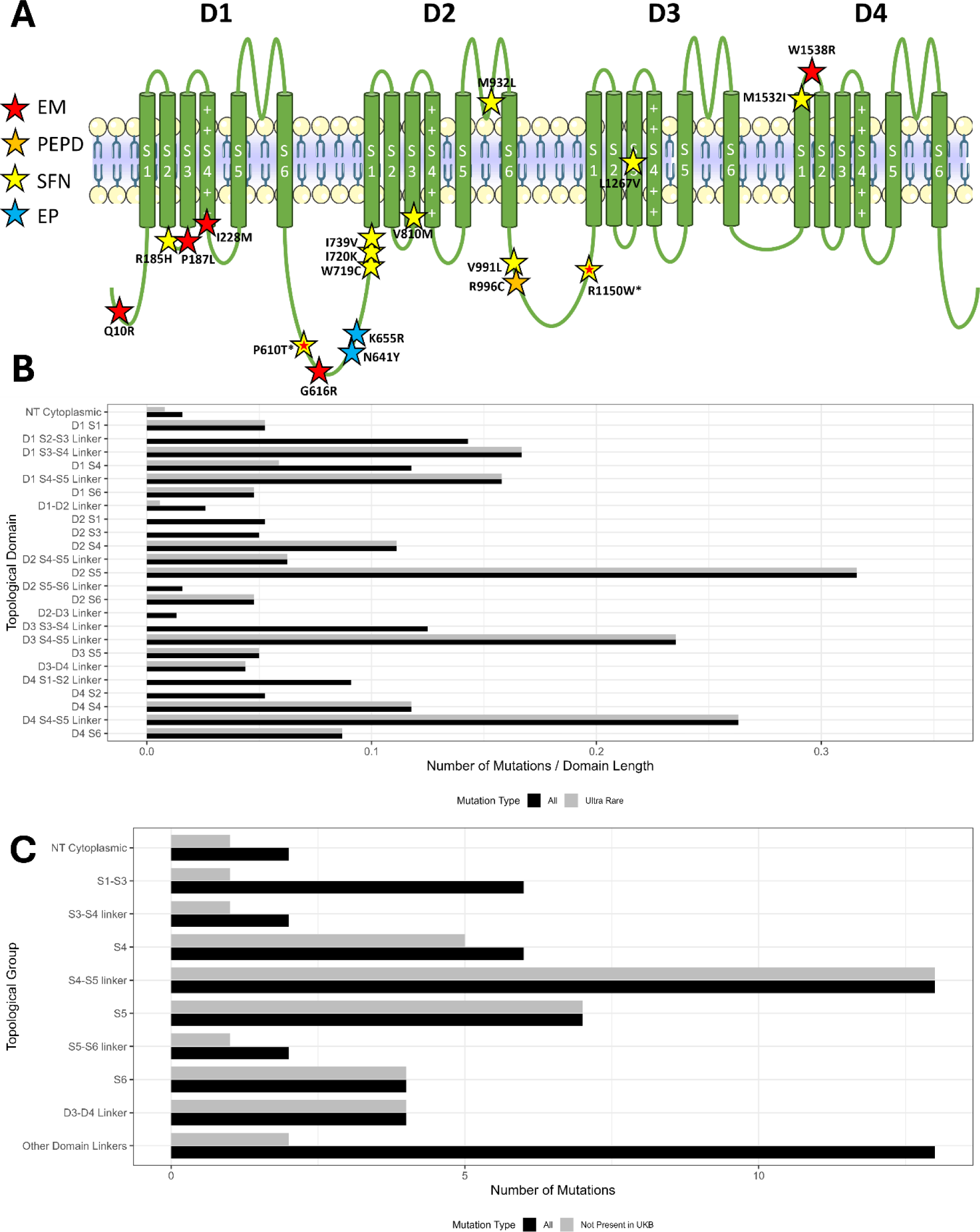
**A.** Schematic of NaV1.7 annotated with mutations present in UK Biobank identified with at least 1 heterozygous carrier. **Red** star = erythromelalgia (EM), **Orange** star = paroxysmal extreme pain disorder (PEPD), **Yellow** star = small-fibre neuropathy (SFN), **Blue** star = epilepsy (EP). * = mutation initially described in erythromelalgia patients but later considered to be small-fibre neuropathy mutation. **B.** Distribution of all know NaV1.7 (*N* = 59) gain-of-function channelopathy mutations (**Black**) across the topological domains of the channel expressed as mutation rate (number of mutations in domain / length of domain in amino acids). Ultra rare mutation group (**Grey**) is the mutations that were not detected in the UK Biobank cohort. Domains are ordered from top to bottom in the order they appear in the NaV1.7 protein. NT = N-Terminus, D = Domain, S = Segment. **C.** Distribution of NaV1.7 gain-of-function channelopathy mutations collapsed into segments across domains in a manner similar to Brunklaus *et al.*, 2022 ^63^. NT = N-Terminus, D = Domain, S = Segment, Other Domain Linkers = D1-D2 + D2-D3 Linkers.

### Many UK Biobank participants carry putative pathogenic *SCN9A* variants

We found 34% of these putative pathogenic *SCN9A* variants, (*20 / 59*) were carried by one or more participants in the UK Biobank (**Figure 1A**, **Table 1**). Most of these *SCN9A* variants (*18 / 20*) had gain-of-function demonstrated in whole-cell patch clamp studies (**Table 1**). These *SCN9A* variants in UK Biobank were causally implicated in erythromelalgia (*N = 8, 40%*), PEPD (*N = 1, 5%*), epilepsy (*N = 2, 10%*) and the remainder were associated with small fibre neuropathy (*N = 9, 45%*) (**Table 1**). In aggregate, a total of *148,901 / 469,779* of the UK Biobank participants (*31.6%*) carried at least 1 of these putatively pathogenic *SCN9A* variants and of these 8,852 were homozygous carriers (**Table 1**).

**Table 1.** Minor allele frequencies, counts, pathogenicity predictions, and intra- and inter-species conservation for the 20 published NaV1.7 channelopathy variants observed in the UK Biobank dataset. **Green =** mutation carried forward for logistic regression. **Yellow** = Mutations in tight linkage disequilibrium and thus not possible to dissect individual mutation contributions (multiple mutation carriers not included in study). **Grey** = mutation removed from further analysis due to rarity. **Red** = mutation removed from further analysis due to commonality. Het = Heterozygous carriers, Hom = Homozygous carriers, UKB MAF = UK Biobank MAF, 1000G = 1000 Genomes Project MAF, gnomAD = Genome Aggregation Database MAF, Ephys = Electrophysiology gain-of-function, AM = AlphaMissense, Inter = Interspecies NaV1.7 sequence alignment score, Intra = Intraspecies human NaV family protein alignment. EM = erythromelalgia, PEPD – Paroxysmal Extreme Pain Disorder, EP – Epilepsy, SFN – Small Fibre Neuropathy. * = originally proposed as an EM mutation, but now considered to be SFN.

| Nav1.7<br>Mutation | Het | Hom | UKB MAF | 1000G<br>MAF | gnomAD<br>MAF | Ephys<br>GoF | Disease | UniProt | ClinVar | AM | EVE | CADD | Inter | Intra |
| --- | --- | --- | --- | --- | --- | --- | --- | --- | --- | --- | --- | --- | --- | --- |
| **Q10R** | 3 | 0 | <0.0001 | - | 0.0001 | Y | EM | Pathogenic | Likely benign | 0.097 | NA | 22.5 | 10 | 1 |
| **R185H** | 1883 | 4 | 0.0020 | 0 | 0.0054 | Y | SFN | Benign | Benign/Likely benign | 0.401 | 0.881 | 27.6 | 10 | 9 |
| **P187L** | 15 | 0 | <0.0001 | - | <0.0001 | N | EM | Uncertain significance | Uncertain significance | 0.732 | 0.892 | 27.6 | 10 | 10 |
| **I228M** | 1381 | 1 | 0.0015 | 0.001 | 0.0008 | Y | EM | Benign | Conflicting interpretations | 0.722 | 0.538 | 17.64 | 10 | 8 |
| **P610T\*** | 24430 | 375 | 0.0275 | 0.0328 | 0.0209 | Y | SFN | Benign | Benign/Likely benign | 0.08 | 0.205 | 17.43 | 10 | 6 |
| **G616R** | 78 | 0 | 0.0001 | - | 0.0001 | Y | EM | Pathogenic | Uncertain significance | 0.437 | 0.293 | 22.6 | 10 | 6 |
| **N641T** | 22 | 0 | <0.0001 | - | <0.0001 | Y | EP | Benign | Conflicting interpretations | 0.172 | 0.218 | 25.8 | 10 | 1 |
| **K655R** | 2519 | 4 | 0.0027 | 0.003 | 0.0017 | Y | EP | Benign | Conflicting interpretations | 0.069 | 0.113 | 12.78 | 10 | 3 |
| **W719C** | 189 | 1 | 0.0002 | 0.001 | 0.0031 | Y | SFN | Benign | Benign/Likely benign | 0.942 | 0.746 | 28.7 | 10 | 10 |
| **I720K** | 256 | 0 | 0.0003 | 0.002 | 0.0002 | Y | SFN | Likely benign | Conflicting interpretations | 0.162 | 0.218 | 23.6 | 10 | 2 |
| **I739V** | 3877 | 7 | 0.0042 | 0.003 | 0.0027 | Y | SFN | Pathogenic | Conflicting interpretations | 0.115 | 0.376 | 22.1 | 10 | 9 |
| **V810M** | 215 | 2 | 0.0002 | 0.001 | 0.0001 | Y | SFN | Benign | Conflicting interpretations | 0.175 | 0.096 | 16.3 | 9 | 3 |
| **M932L** | 2644 | 22 | 0.0029 | 0 | 0.0147 | Y | SFN | NA | Benign/Likely benign | 0.13 | 0.155 | 19.28 | 10 | 6 |
| **V991L** | 2642 | 21 | 0.0029 | 0 | 0.0145 | Y | EM | Pathogenic | Conflicting interpretations | 0.112 | 0.163 | 5.961 | 10 | 7 |
| **R996C** | 15 | 0 | <0.0001 | 0 | <0.0001 | N | PEPD | Pathogenic | Conflicting interpretations | 0.106 | 0.179 | 16.58 | 9 | 3 |
| **R1150W\*** | 108583 | 8405 | 0.1540 | 0.1128 | 0.1215 | Y | EM | Benign | NA | NA | NA | NA | 10 | 4 |
| **L1267V** | 2396 | 4 | 0.0026 | 0.002 | 0.0017 | Y | SFN | Pathogenic | Conflicting interpretations | 0.144 | 0.281 | 24.3 | 10 | 8 |
| **M1532I** | 4 | 0 | <0.0001 | - | <0.0001 | Y | SFN | Uncertain significance | Uncertain significance | 0.31 | 0.189 | 7.615 | 8 | 6 |
| **W1538R** | 2245 | 6 | 0.0024 | 0 | 0.0019 | Y | EM | Benign | Conflicting interpretations | 0.299 | 0.162 | 21.6 | 10 | 4 |

### Frequency of occurrence of NaV1.7 mutations

The SCN9A allele frequencies calculated from the UK Biobank whole exome sequencing dataset were similar to those previously reported in the 1000 Genomes Project and gnomAD 4.1.0 excluding UK Biobank data (**Table 1**). A total of 18 of the identified variants in UK Biobank were considered to be rare with a minor allele frequency (MAF) < 0.01^51^. The remaining two variants that cause the NaV1.7 P610T and R1150W mutations had MAFs of 0.028 and 0.15, respectively. The commonality of the R1150W variant, found in *116,988* participants, suggests it is extremely unlikely be responsible for a highly penetrant pain phenotype. A sensitivity analysis supported this conclusion as a combined group of hetero- and homozygous carriers of R1150W do not have an increased risk of chronic pain (*Fisher’s Exact Test, OR = 1.00, 95% CI = 0.98-1.01, P = 0.5*). The commonality of R1150W also caused potential issues for downstream analysis, as participants carrying more than one mutation were excluded meaning those participants carrying other variants would have been excluded – substantially reducing the numbers in those smaller allele groups. Therefore, R1150W carrier status was not further considered during this analysis.

For the binomial regression analysis, we included only those NaV1.7 mutations that had ≥ 50 carriers in UK Biobank which led to the exclusion of a further 6 groups leaving 13 variants for individual regression analysis (**Table 1**). The detailed results for 3 of the NaV1.7 mutations, selected as they had some of the strongest supporting evidence for pathogenicity in the prior literature, are explored below.

### There is no evidence for a pain phenotype in carriers of NaV1.7 I228M

NaV1.7^I228M^ is the most extensively studied painful channelopathy mutation. It was initially reported in 3 patients across 2 families ^52^ and each had a presentation consistent with small fibre neuropathy – one with features characteristic of erythromelalgia. An independent research group reported this mutation in another erythromelalgia patient^53^.

Microneurography showed that this patient had spontaneous activity in their C-fibres and an abnormal pattern of activity-dependent slowing^21^. Whole-cell voltage clamp in HEK293 cells transfected with NaV1.7^I228M^ revealed a significant depolarising shift of 6.8 mV in slow inactivation of the sodium conductance^52^. Current clamp recordings in cultured rat DRG neurons transfected with NaV1.7^I228M^ identified a 4.8 mV depolarising shift in resting membrane potential and spontaneous firing in 29% of neurons. Similar recordings in cultured transfected trigeminal ganglia (TG) neurons revealed an 8.5 mV depolarising shift in resting membrane potential and a 36% reduction in the threshold for action potential discharge. Cultured DRG neurons expressing NaV1.7^I228M^ have significantly reduced neurite outgrowth, mitochondrial size and density^54,55^. *In vivo*, a genetically modified mouse homozygous for NaV1.7^I228M^ develops skin lesions, an insensitivity to noxious hot and cold, and to the pruritogen chloroquine accompanied by a pronounced decrease in the expression of marker genes for C-fibre low-threshold mechanoreceptive neurons ^56^. The introduction of the NaV1.7^I228M^ variant to zebrafish has been proposed as a model of small-fibre neuropathy^57^.

The amino acid mutated in I228 is a highly conserved residue across species (*10 / 10*) and within the human NaV family (*8 / 10*) (**Table 1**). ClinVar reported “conflicting interpretations of pathogenicity” and UniProt categorised it as “benign” (**Table 1**). Both AlphaMissense and CADD predict I228M to be pathogenic, whereas EVE reports it as ambiguous (**Table 1**). Considering these predictions alongside the evidence base for the functional effects of this mutation, it was anticipated that I228M would be associated with a penetrant pain phenotype.

There are *1381* heterozygous carriers and one homozygous carrier of the I228M mutation in UK Biobank, with a MAF of *0.001*. The I228M mutation was not significantly associated with chronic pain (*OR = 0.97, 95% CI = 0.87-1.08, P = 0.95, N = 1332*) or neuropathic pain (*OR = 1.04, 95% CI = 0.73-1.49, P = 0.94, N = 202*) (**Figure 2A**). Similarly, there was no significantly increased prevalence of prescription of analgesic or anti-neuropathic pain medications (**Figure 2A**).

**Figure 2.**
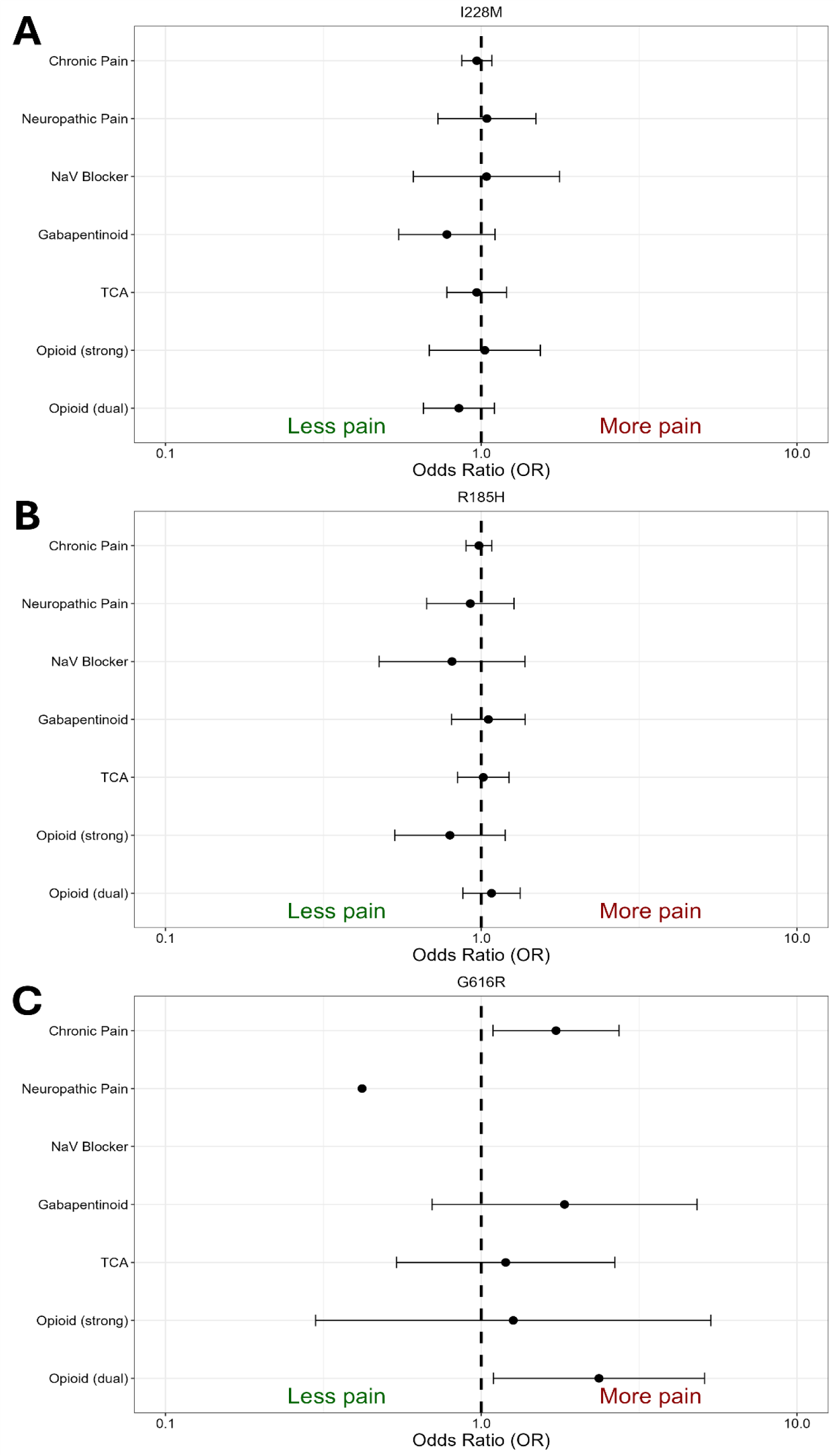
NaV1.7 I228M **(A)**, NaV1.7 R185H **(B)**, and NaV1.7 G616R **(C)** are not associated with an increased risk of any pain phenotype. Odds ratios for each phenotype are presented with their 95% confidence intervals.

### There is no evidence for a pain phenotype in carriers of NaV1.7 R185H

The R185H mutation was first reported in two unrelated patients with small fibre neuropathy^58^. The same mutation has also been identified in 2 siblings with epilepsy and febrile seizures^59^. Whole-cell patch clamp in cultured transfected mouse DRG neurons revealed a 35% reduction in the current threshold for action potential discharge perhaps related to an increase in resurgent currents^60^. No other abnormalities in the biophysical properties of the mutant NaV1.7 channel were observed. A *SCN9A^R185H^* genetically modified mouse line has been generated which exhibits thermal and mechanical hyperalgesia ^61^ and clonidine elicited a conditional place preference suggesting relief of an ongoing spontaneous pain phenotype.

The NaV1.7 R185 residue was highly conserved across species (*10 / 10*) and within the human NaV protein family (*9 / 10*) (**Table 1**). UniProt and ClinVar categorised it as benign (**Table 1**). The EVE and CADD models predicted R185H to be pathogenic whereas AlphaMissense reported it as ambiguous (**Table 1**).

The R185H mutation has 1,883 heterozygous and 4 homozygous carriers in the UK Biobank (MAF 0.002). NaV1.7^R185H^ was not associated with an increased risk of chronic pain (*OR = 0.98, 95% CI = 0.90-1.08, P = 0.95, N = 1821*) or neuropathic pain (OR = 0.92, 95% CI = 0.67-1.27, *P* = 0.95*, N = 271*) (**Figure 2B**). Similarly, NaV1.7^R185H^ was not associated with a significantly increased prevalence of prescriptions of analgesics or anti-neuropathic pain medications (**Figure 2B**).

### There is no evidence for a pain phenotype in carriers of NaV1.7 G616R

NaV1.7^G616R^ was initially observed in 4 erythromelalgia patients from the same family, across 3 generations, segregating with the phenotype in 4 of the 5 mutation carriers^62^. The age of onset for this family varied from 10 – 24 years, which may explain why a 17-year-old carrier had not yet developed erythromelalgia. In whole-cell patch clamp of transfected cultured mouse DRG neurons, NaV1.7^G616R^ currents had a 5.7 mV depolarising shift in inactivation and was associated with spontaneous firing in 33% of neurons^62^.

The G616 residue was highly conserved across species (10 / 10) and was moderately conserved within the human NaV protein family (6 / 10) (**Table 1**). UniProt categorised it as pathogenic, whereas ClinVar annotates it as uncertain (**Table 1**). It is predicted to be pathogenic by CADD, ambiguous by AlphaMissense, and benign by EVE (**Table 1**).

With only 78 heterozygous carriers in the UK Biobank cohort, G616R is very rare (MAF 0.0001). It was not significantly associated with chronic pain (*OR = 1.73, 95% CI = 1.09-2.73, P = 0.46, N = 74*) nor with neuropathic pain (*OR = 0.41, 95% CI = 0.05-3.25, P = 0.85, N = 12*) (**Figure 2C**). It was not associated with a significantly increased prevalence of prescriptions of analgesics or anti-neuropathic pain medications (**Figure 2C**).

### There is no evidence for a pain phenotype in carriers of any of the other NaV1.7 mutations

Similar analysis was applied to the other 8 NaV1.7 mutations suggested to be pathogenic from the literature search and with > 50 carriers in UK Biobank. These included mutations reported to cause EM (2) and SFN (7). The carriers of these variants showed no evidence of a significantly increased incidence of chronic pain, prescription analgesics, or anti-neuropathic pain medications (**Table 2**).

**Table 2.** Summary of Odds ratios [±95% CI] values coloured by FDR adjusted *P* value (>0.05).

| **Mutation** | **Chronic Pain** | **Neuropathic Pain** | **NaV Blocker** | **TCA** | **Gabapentinoid** | **Opioid (strong)** | **Opioid (dual)** |
| --- | --- | --- | --- | --- | --- | --- | --- |
| **R185H** | **0.98**<br>[0.90, 1.08] | **0.92**<br>[0.67, 1.27] | **0.81**<br>[0.47, 1.37] | **1.01**<br>[0.84, 1.22] | **1.05**<br>[0.80, 1.38] | **0.80**<br>[0.53, 1.19] | **1.08**<br>[0.87, 1.33] |
| **I228M** | **0.97**<br>[0.87, 1.08] | **1.04**<br>[0.73, 1.49] | **1.04**<br>[0.61, 1.77] | **0.97**<br>[0.78, 1.20] | **0.78**<br>[0.55, 1.11] | **1.03**<br>[0.68, 1.54] | **0.85**<br>[0.66, 1.10] |
| **P610T** | **1.00**<br>[0.97, 1.03] | **0.96**<br>[0.88, 1.05] | **1.01**<br>[0.88, 1.15] | **0.97**<br>[0.92, 1.02] | **0.92**<br>[0.85, 1.00] | **0.93**<br>[0.84, 1.03] | **0.96**<br>[0.90, 1.02] |
| **G616R** | **1.72**<br>[1.09, 2.73] | **0.42**<br>[0.05, 3.25] | **0.00**<br>[0.00, Inf] | **1.19**<br>[0.54, 2.65] | **1.84**<br>[0.70, 4.83] | **1.26**<br>[0.30, 5.34] | **2.36**<br>[1.09, 5.09] |
| **K655R** | **0.96**<br>[0.88, 1.04] | **1.06**<br>[0.83, 1.36] | **1.26**<br>[0.88, 1.81] | **0.98**<br>[0.84, 1.15] | **0.99**<br>[0.79, 1.25] | **0.96**<br>[0.70, 1.30] | **1.00**<br>[0.83, 1.20] |
| **W719C** | **0.66**<br>[0.49, 0.89] | **0.81**<br>[0.17, 3.86] | **0.00**<br>[0.00, Inf] | **0.56**<br>[0.26, 1.21] | **0.58**<br>[0.18, 1.89] | **0.83**<br>[0.20, 3.47] | **0.61**<br>[0.25, 1.44] |
| **I720K** | **0.87**<br>[0.67, 1.12] | **0.72**<br>[0.30, 1.69] | **1.73**<br>[0.63, 4.73] | **0.66**<br>[0.37, 1.16] | **1.05**<br>[0.51, 2.19] | **0.92**<br>[0.34, 2.52] | **1.29**<br>[0.75, 2.21] |
| **I739V** | **1.04**<br>[0.97, 1.11] | **0.96**<br>[0.78, 1.18] | **0.70**<br>[0.48, 1.03] | **1.15**<br>[1.01, 1.30] | **1.07**<br>[0.89, 1.28] | **1.25**<br>[1.01, 1.56] | **1.13**<br>[0.98, 1.30] |
| **V810M** | **1.08**<br>[0.82, 1.42] | **1.10**<br>[0.51, 2.39] | **1.63**<br>[0.51, 5.20] | **0.92**<br>[0.51, 1.65] | **0.95**<br>[0.41, 2.20] | **0.83**<br>[0.26, 2.65] | **1.42**<br>[0.79, 2.54] |
| **L1267V** | **1.02**<br>[0.94, 1.11] | **0.87**<br>[0.65, 1.16] | **0.72**<br>[0.45, 1.15] | **1.13**<br>[0.96, 1.32] | **0.97**<br>[0.77, 1.23] | **1.05**<br>[0.78, 1.41] | **1.12**<br>[0.94, 1.33] |
| **W1538R** | **0.96**<br>[0.78, 1.17] | **1.77**<br>[1.05, 2.98] | **1.23**<br>[0.50, 3.00] | **0.87**<br>[0.58, 1.30] | **1.58**<br>[0.99, 2.52] | **0.67**<br>[0.27, 1.63] | **0.81**<br>[0.51, 1.29] |

We aggregated the 5 rare NaV1.7 mutations (Q10R, P187L, N641Y, R996C, M1532I) that had carriers in UK Biobank (total of 58 carriers of mutations associated with EM (2), PEPD (1), SFN (1) and EP (1)) into a composite group and again found no evidence of an increased incidence of chronic pain (*OR = 1.03, 95% CI = 0.61-1.74, P = 0.90, N = 58*) (**Supplementary Table 6**). As a further sensitivity analysis we also aggregated all NaV1.7 mutation carriers (including the rare mutations) together with participants carrying multiple NaV1.7 mutations and found no evidence for an increased risk of chronic pain (*OR = 0.99, 95% CI = 0.97 -1.02, P = 0.62, N = 40,236*) (**Supplementary Table 7**).

A grouped analysis of the pathogenicity predictions for the variants observed in the UK Biobank dataset compared to those which were absent showed that the ultra-rare NaV1.7 variants had significantly greater predicted risk of pathogenicity (**Figure 3**, assessed with CADD, AlphaMissense and EVE). Re-plotting the distribution of these ultra-rare NaV1.7 variants showed that they were concentrated in the functionally important and conserved regions of the sodium channel protein (like the S4, S5, S6 and D3-D4 linkers) with a winnowing out of the UK Biobank NaV1.7 variants that were mostly found in non-conserved regions like the D1-D2 and D2-D3 domain linkers (**Figure 1B**). Further collapsing the mutations in a similar method to Brunklaus et al., 2022^63^ supported this conclusion (**Figure 1C**).

**Figure 3.**
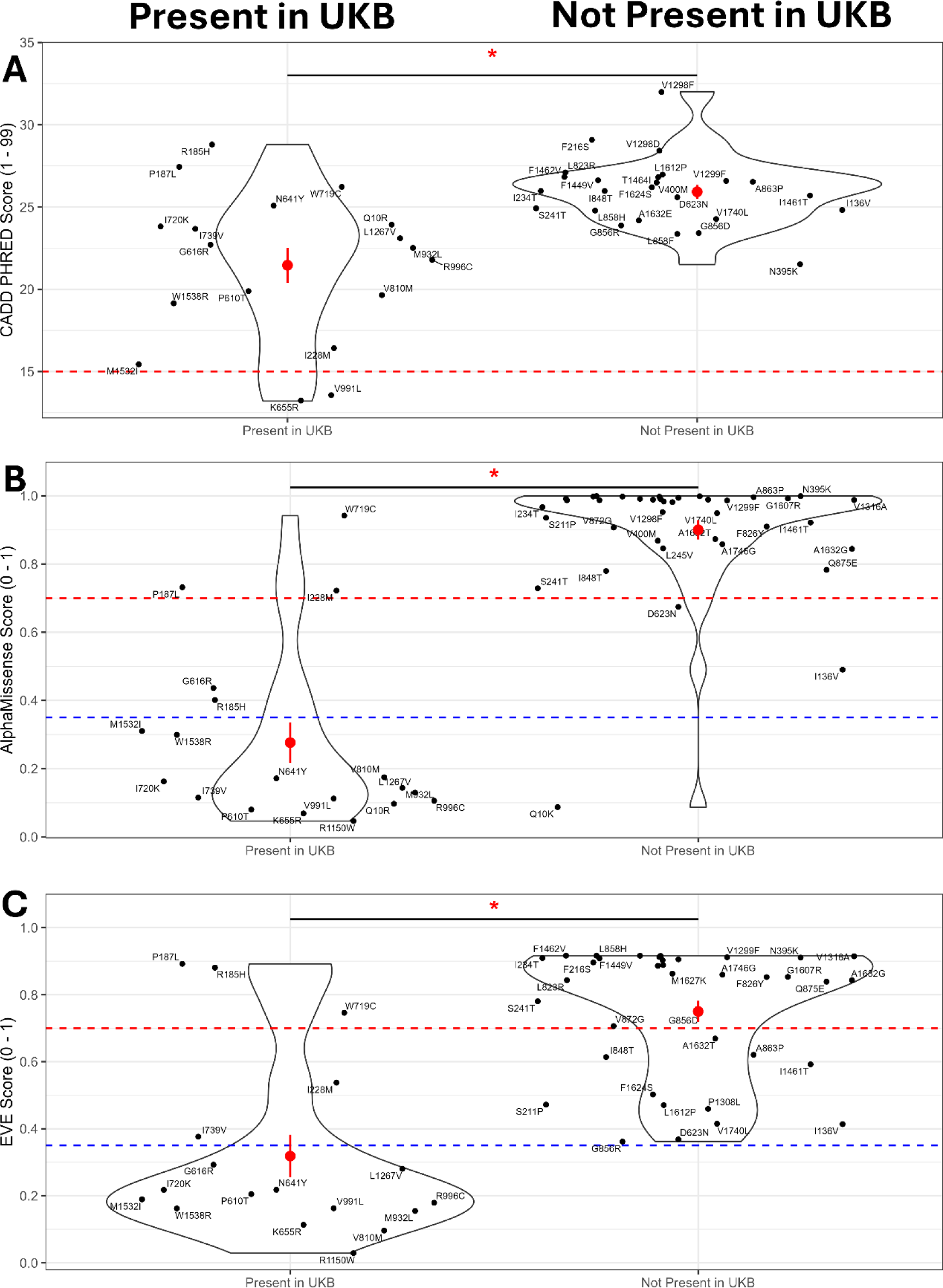
Violin plots of pathogenicity predictions from 3 machine learning models, CADD **(A)**, AlphaMissense **(B)**, and EVE **(C)**, grouped by those observed in UK Biobank dataset or absent (Ultra Rare). All mutations observed in UK Biobank had significantly lower average pathogenicity scores than the Ultra Rare group (CADD: *W* = 81, *P* = 0.0004; AlphaMissense: *W* = 35, *P* = 2.471 × 10^-10^; EVE: *W* = 70, *P* = 2.903 × 10^-7^). Red dot and line = mean ± standard error. The dashed red line = pathogenic threshold, and the dashed blue line = ambiguous threshold.

### There is no evidence for a pain phenotype in carriers of (suspected) Nav1.7 LoF mutations

139 suspected loss-of-function SCN9A variants were identified using VEP, where 719 heterozygous carriers were detected in UK Biobank. These suspected heterozygous SCN9A loss-of-function variants were not found to be significantly associated with an altered risk of chronic pain (OR 0.96[0.82-1.11], P=0.99) nor neuropathic pain (OR 1.15[0.68-1.84], P=0.99) (figure 4). Similarly, carriers of these variants were not significantly associated with changes to analgesic prescription, including gabapentinoids (OR 1.05[0.68-1.54], P=0.99), Nav-channel blockers (OR 0.86[0.34-1.76], P=0.99), dual-mechanism opioids (OR 0.70[0.48-0.99], P=0.49), strong opioids (OR 1.12[0.63-1.83], P=0.99), and tricyclic antidepressants (OR 1.01[0.77-1.31], P=0.99) (figure 4).

### There is no evidence for a pain phenotype in carriers of Nav1.8 GoF mutations

Using the search strategy, 5 SCN10A gain-of-function variants were identified, with 4 present in the UK Biobank, comprising 1,104 heterozygous carriers. 3 of these variants were isolated from patients with small-fibre neuropathy, whilst 1 was identified in a patient with erythromelalgia, all being heterozygous (table 5). Carriers of these gain-of-function variants were not found to be associated with a higher risk of either chronic (OR 0.96[0.85-1.08], P=0.94) or neuropathic pain (OR 1.02[0.67-1.49], P=0.96), (figure 6). There was no significant association found between carriers and changes to analgesic prescription, including gabapentinoids (OR 0.94[0.67-1.28], P=0.95), Nav-channel blockers (OR 1.18[0.65-1.96], P=0.95), dual-mechanism opioids (OR 0.99[0.77-1.26], P=0.96), strong opioids (OR 0.77[0.44-1.23], P=0.84), and tricyclic antidepressants (OR 1.15[0.94-1.40], P=0.84) (figure 6).

### There is no evidence for a pain phenotype in carriers of Nav1.8 LoF mutations

167 SCN10A loss-of-function variants were identified using VEP, with 1,308 heterozygous carriers found in the UK Biobank. Suspected loss-of-function in SCN10A was not found to be significantly associated with an altered risk of chronic pain (OR 1.13[1.01-1.26], P=0.67) or neuropathic pain (OR 0.82[0.54-1.20], P=0.84) (figure 7). There was no significant association found between carriers and changes to analgesic prescription, including gabapentinoids (OR 0.99[0.72-1.33], P=0.96), Nav-channel blockers (OR 1.08[0.61-1.75], P=0.95), dual-mechanism opioids (OR 1.22[0.98-1.50], P=0.67), strong opioids (OR 574 1.18[0.79-1.69], P=0.86), and tricyclic antidepressants (OR 1.16[0.96-1.39], P=0.84) (figure 575 7).

### There is no evidence for a pain phenotype in carriers of Nav1.9 GoF mutations

Using the search strategy, 9 gain-of-function SCN11A variants were identified, 6 of which were present in the UK Biobank, comprising 476 heterozygous carriers (table 6). Of these, 3 were identified in patients with FEPS, and 3 in patients with small-fibre neuropathy, all heterozygous. SCN11A gain-of-function was not found to be significantly associated with changes in chronic (OR 1.00[0.84-1.20], P=0.99) or neuropathic pain (OR 1.26[0.72-2.10], P=0.97) (figure 11). There were no significant associations found with carriers and changes to analgesic prescriptions, including gabapentinoids (OR 1.31[0.82-1.98], P=0.86), Nav-channel blockers (OR 0.80[0.39-1.46], P=0.99), dual-mechanism opioids (OR 1.00[0.68-1.43], P=0.99), strong opioids (OR 1.22[0.63-2.12], P=0.99) or tricyclic antidepressants (OR 1.53[1.15-1.98], P=0.21) (figure 11). SCN11A gain-of-function variants possessing >50 carriers were analysed individually.

### There is no evidence for a pain phenotype in carriers of Nav1.9 LoF mutations

158 suspected SCN11A loss-of-function mutations were identified using VEP, comprising 1161 heterozygous carriers and 8 compound heterozygous carriers. These loss-of-function variants were not found to be significantly associated with changes in chronic (OR 0.90[0.80-1.02], P=0.98) or neuropathic pain (OR 1.25[0.87-1.76], P=0.87) (figure 14). There was also no significant association with changes in analgesic prescription, including gabapentinoids (OR 1.29[1.11-2.92], P=0.19), Nav-channel blockers (OR 0.80[0.24-2.58], P=0.99), dual-mechanism opioids (OR 1.12[0.88-1.41], P=0.98), strong opioids (OR 1.03[0.87-1.31], P=0.73), or tricyclic antidepressants (OR 1.08[0.87-1.32], P=0.44) (figure 14).

## Discussion

By taking advantage of the scale of the UK Biobank cohort and the release of the whole exome sequencing data we found that NaV1.7 mutations reported to cause erythromelalgia, PEPD, or small-fibre neuropathy were carried by almost 35% of this population sample. Considering that almost all of these mutations had gain-of-function electrophysiological evidence, and in some cases even human *in vivo* evidence showing spontaneous C-fibre nociceptor firing, a highly penetrant pain phenotype was expected. No evidence of a highly penetrant pain phenotype was found for the NaV1.7 I228M, G616R, or W1538R mutations which have all been reported as causal for erythromelalgia^10,52,53,62^, and which were therefore expected to be strongly associated with pain. Similarly, there were no associations with pain for any of the small-fibre neuropathy mutations (R185H, P610T, W719C, I720K, I739V, V810M, M932L, and L1267V). It is also notable that the remaining two thirds of the putative pathogenic variants, including all of the PEPD mutations, were very rare and had insufficient (or in most cases zero) carriers in UK Biobank for individual analysis – potentially consistent with them being pathogenic.

Following the original linkage of *SCN9A* variants to erythromelalgia^8^, there has been a steady increase in the number novel mutations reported in patients. Early studies that identified the first erythromelalgia mutations used Sanger sequencing of the exons and flanking regions of the *SCN9A* gene to identify polymorphisms^8,64^. Following the identification of a potential novel pathogenic mutation, the next step is to assess sequence conservation across species to determine if it is likely to be essential for function^65^. It is then necessary to determine whether the variant responsible for the mutation is rare^66^. A minor allele frequency of less than 1% is typically used for filtering potential candidate causal variants. However, in practice, the minor allele frequency of autosomal dominant Mendelian disease variants is usually under 0.01%^67^. For some erythromelalgia studies, small ethnicity-matched control cohorts of 25 – 100 people had their *SCN9A* gene sequenced to determine if a variant was rare^64^. This degree of sequence conservation and variant rarity, combined with familial co-segregation of the mutation and the phenotype, has been used to support the case for pathogenicity^8,68^. Several more recent studies have sequenced panels of 20 – 30 candidate pain genes^53,69^. However, these approaches still suffer to a degree from the same issue as sequencing *SCN9A* alone – one of confirmation bias by limiting the search to already known candidate genes. Advances in sequencing technology have made it more cost-efficient to sequence a whole exome than to carry out targeted sequencing – and this reduces the risk of false positive associations by allowing the influence of variants in other genes to be explored. Of the 59 reported gain-of-function pathogenic *SCN9A* variants, only 2 (3%) were identified in studies that used whole exome sequencing.

As a consequence of the methods historically employed in the identification of candidates, it is likely that some *SCN9A* variants may have been mislabelled as pathogenic and also that causal variants in other genes have likely been missed for some of these cases. In two previous erythromelalgia cohort studies, only 15% (2 / 7) and 28% (7 / 48) of patients had a rare *SCN9A* protein coding region variant, and all of those NaV1.7 mutations observed within the cohorts had been previously reported in other erythromelalgia studies suggesting that we are approaching saturation in the discovery of new causal variants^21,53^. Given these findings, it is increasingly likely that the majority of cases of erythromelalgia are not due to gain-of-function NaV1.7 mutations. It is also relevant that there has been a report of erythromelalgia in a patient with a variant in the *SCN10A* gene that caused the NaV1.8 M650K missense mutation ^70^ and this has been shown to confer electrophysiological gain of function in vitro and microneurography in the patient showed spontaneous activity in nociceptors. This case supports the principle that erythromelalgia may be due to mutations in proteins other than NaV1.7.

There is a variability in the phenotypes of the erythromelalgia patients, including between those carrying identical *SCN9A* mutations and even within members of the same family^11,52^. This is the also the case for other genetic conditions like cystic fibrosis, where it is accepted that part of the phenotypic variability is due to mutations in modifier genes that exacerbate or partially relieve symptoms^71^. Of particular relevance to this question of variability is a study of inherited erythromelalgia where the daughter had substantially worse pain phenotype than her mother. They both carried the same NaV1.7 F1449V mutation (not present in UK Biobank). Exome sequencing revealed that the mother was heterozygous for a potassium channel variant *in KCNQ3*. Whole-cell voltage clamp analysis in HEK293 cells revealed that the Kv7.3 D755N mutation produced an M current (inwardly rectifying potassium current) that exhibited significantly less time-dependent conductance reduction than the wild-type. The M current contributes to setting the action potential threshold in cells leading to the suggestion that this may have compensated partially for the *SCN9A* mutant’s increase of nociceptor excitability and led to a less severe pain phenotype^72^. It is reasonable to assume that there are likely to be modulator genes involved in the development of the erythromelalgia phenotype, acting to either worsen or attenuate the pain severity, and thus alter the likelihood of being diagnosed with the disease. Another possibility that should be considered is the contribution of environmental triggers to diseases like erythromelalgia and PEPD. Relatively little is known about the environmental interactions that can influence these pain diseases. It is reasonable to expect that there may be unknown environmental or gene-environment interactions at play^52^.

While some NaV1.7 mutation carriers may possess protective gene variants that mask the pain phenotype, it is unlikely that this is sufficient to explain the lack of increased risk of chronic pain in participants carrying mutations that should make their sensory neurons hyperexcitable. It appears that some of these mutations are not pathogenic in humans *in vivo,* and perhaps their gain-of-function is only present in a reduced *in vitro* preparation. This could be due to a relative overexpression of the NaV1.7 channel, the lack of normal regulatory interacting proteins, abnormal distribution of proteins within cell lines and alterations to cellular phenotype from the culture conditions or differentiation protocol. We note that only one electrophysiological study from those reviewed presented negative findings from functional testing of potential erythromelalgia mutations^73^ perhaps suggesting the presence of a positive publication bias. The typical discovery sequence for a potentially pathogenic NaV1.7 variant from clinical phenotyping a rare pain patient with unusual features, through family pedigree and genetic association with targeted sequencing will usually have preceded any electrophysiological studies which are then conducted in the expectation that there should be a gain of function in some aspect of channel biophysics as a last piece in the evidence puzzle. These electrophysiological findings are seldom biophysically modelled within a “nociceptor” to see whether the change of behaviour could account for increased excitability or spontaneous activity. Furthermore, these variants are rarely placed back into the in vivo context and where that has been done, for example for the EM I228M mutation, in mouse ^56,74^ or zebrafish ^57^ models the findings have often not recapitulated the erythromelalgia phenotype. Perhaps these NaV1.7 mutations do not cause the same gain-of-function *in vivo* that they do *in vitro*, or maybe they cause hyperexcitability of DRG neurons, but this is insufficient to cause pain. The potential for inaccurate assignation of pathogenicity for the SFN variants is likely greater and accordingly a higher proportion (88%) of these variants are present in UK Biobank (8 / 9) and none showed any evidence of an association with a pain phenotype. This likely reflects the weaker association for the SFN NaV1.7 mutations to the underlying pathology with no family pedigree indicating heritability and no significant linkage to the presence of a painful neuropathy. It seems that the field will need to be more critical in its appraisal of in vitro electrophysiological findings which have hitherto been regarded as “gold standard” pieces of evidence for identifying the gain of function in pathogenic NaV1.7 mutations.

The NaV1.7 mutations that were either not observed in the UK Biobank dataset at all or were excluded from analysis because they had < 50 carriers, are more likely to be pathogenic due to their rarity. Of the 59 SCN9A variants reported in patients, 39 were so rare that they were not observed in the UK Biobank dataset, nor can they be found in the gnomAD database (v4.1.0). There was a substantial and statistically significant difference in the average CADD, AlphaMissense, and EVE pathogenicity scores of mutations observed in UK Biobank versus those that were ultra rare and thus not observed in the dataset (although only EVE does not use MAF as either a model feature (CADD) or for fine-tuning (AlphaMissense))^35,75^. Another method to triangulate the predictions of pathogenicity is through the comparative analysis of paralogues of each variant in the other human NaVs where there is strong sequence conservation in the functionally important domains^41^. This showed that 37% of the ultra-rare variants not present in the UK Biobank dataset had paralogues with documented electrophysiological evidence of altered function whereas none of the variants present in UK Biobank had any corroborating evidence (https://scn-viewer.broadinstitute.org/, **Supplementary Table 5**). Similarly, almost all (94%) of the pathogenic NaV1.7 mutations that were not observed in UK Biobank had paralogues that were associated with diseases like epilepsy, long QT syndrome, and myotonia whereas only a minority (32%) of the NaV1.7 mutations observed in UK Biobank had paralogues associated with diseases. This can also be seen in the relative positions of the rare variants within the protein structure which are concentrated in the most conserved and functionally important domains such as the S4 voltage sensor, the fast inactivation gate in D3-D4 linker and in the pore domains S5 and S6 unlike that seen in the commoner variants.

The lack of evidence for a high penetrance pain phenotype is perhaps more surprising in gain-of-function mutations in Nav1.8 and 1.9, since the patients from whom these variants were identified exhibited painful neuropathies despite being heterozygous for the mutant gene. It would therefore be expected that these painful phenotypes would be mirrored in other heterozygous carriers of these gain-of-function mutations, however, these were not identified. One potential explanation for this is the involvement of modulator genetic or epigenetic factors, which could have an influence on the outcome of these gain-of-function mutations, perhaps suppressing their effect, and preventing a change in phenotype. Another, perhaps more convincing explanation, could lie in the experimental methods used to identify these variants in the first place. Of the 11 gain-of-function variants identified across Nav1.8 and 1.9, 7 (64%) had only sequenced and examined SCN10A, SCN11A and SCN12A. This is unlike the remaining variants, 3 were identified from whole-exome sequencing, and one from sequencing 29 candidate genes known to be related to nociception. Given the small number of genes examined, it is difficult conclude these genes to be causative for the painful neuropathies seen in their patients, especially given the idiopathic nature of diseases such as small-fibre neuropathy. Utilising methods such as these could lead to an over-ascription of causative variants to painful neuropathy, without sufficient knowledge to assume causality. This uncertainty may explain the lack of painful phenotype seen amongst the carriers within the UK Biobank.

No evidence of a highly penetrant pain phenotype was seen in loss-of-function mutations in Nav1.7. Little research has been undertaken into heterozygous loss-of-function in Nav1.7, so these findings are novel, and are consistent with the lack of symptoms seen in heterozygous carriers. No evidence of a highly penetrant pain phenotype was found in suspected Nav1.8 or 1.9 loss-of-function variants either. This is expected, due to the asymptomatic nature of heterozygous Nav1.7 loss-of-function. One proposed explanation for this is that heterozygous loss-of-function of Nav1.7, 1.8 or 1.9 does not reduce protein expression enough to lead to haploinsufficiency – where a single functional copy of a gene is insufficient to maintain a normal phenotype. The remaining normal gene expresses enough functional protein to maintain a normal phenotype, thus creating a recessive pattern of inheritance. This could result from a level of redundancy within VGSC expression, or perhaps homeostatic mechanisms upregulate expression of the functional gene in response to the mutation. Some evidence from Nav1.7-selective blockers has suggested that >80% channel blockade is required to elicit an analgesic effect, a level much greater than the reduction seen in heterozygous loss-of-function.

One potential limitation of the use of UK Biobank for this study is that it is a volunteer population cohort that recruited ∼6% of the 9 million approached and this led to a known bias towards healthier, more affluent and white population limiting its representativeness of the UK population as a whole^76^. Importantly in this context, the age at recruitment was 37-73, whereas erythromelalgia typically first presents in childhood and PEPD in infancy. Although onset age varies between and within mutation groups, and there have been reports of late adult onset erythromelalgia^77^. Nonetheless, this should have reduced the number of patients with inherited pain conditions. Such an effect would have been expected to reduce the prevalence of NaV1.7 mutations that actually cause inherited painful conditions. The comparison with genomes and exomes sequences from other populations (in gnomAD v4.1.0 non-UKB) is therefore pertinent as the MAFs for these variants is similar. In addition, this healthy volunteer recruitment bias cannot explain how common many of the NaV1.7 variants are in UK Biobank nor their lack of effect on the pain phenotypes.

The depth of prescription data varies, but some participants have data going back to the 1980s. The prescription-based phenotypes in this study only considered whether a participant had ever been prescribed a drug in each class. While the average number of prescriptions was considered as an outcome measure, there were inherent biases with this approach, as some patients had a large number of prescriptions, which disproportionately affected the population estimate. Further analysis of the prescription data could have been justified, for example, considering the dose or participants that have had a number of prescriptions greater than a threshold in a given period, but the lack of signal in the binarized prescribing data allied to the lack of a pain phenotype argued against the value of further exploration.

From a clinical diagnostic perspective, the Association for Clinical Genomic Science recommends using family history, primary literature, gnomAD minor allele frequency, and public databases such as Ensembl and ClinVar to determine pathogenicity^78^. The findings presented here show that many NaV1.7 variants annotated as pathogenic for pain in these databases do not have supporting evidence from the UK Biobank. Familial disease segregation of a rare mutation with strong evolutionary conservation is insufficient to confirm pathogenicity. While *in vitro* functional evidence improves the rationale for attributing pathogenicity to a mutation, this research has highlighted discordance between *in vitro* functional evidence and the lack of an increased prevalence of chronic pain observed in carriers of these mutations in the UK Biobank cohort.

The long story of NaV1.7 as an analgesic target has yet to yield a clinical therapeutic. This is not necessarily because it is an inappropriate target but in part because it has been hard to target. The phase II and III clinical trials of selective small molecule antagonists of NaV1.7 appear to have resulted in them being discontinued^79^. The most compelling evidence for NaV1.7 as an analgesic target is from human genetics, which, to date, has been stronger than any other “pain gene”. It is a gene with bidirectional phenotypic effects – it can cause pain by gain-of-function and insensitivity to pain by loss-of-function^24^. Targets whose genes display this phenomenon are 3.8-fold more likely to be approved as therapeutic agents in humans^80^. Nonetheless the evidence presented here raises caveats about the pathogenicity of a substantial proportion of these gene variants. This may also relate to why the use of the existing non-selective NaV blockers has not been very successful in the treatment of erythromelalgia which is often refractory to current therapeutics^81^.

Nearly all previous NaV1.7 mutations have been identified by targeted sequencing of the *SCN9A* gene in patients. In this study, identifying carriers of previously reported pathogenic mutations and testing to see if carrier groups have a highly penetrant pain phenotype has allowed us to flip the question asked in case studies - “Does this monogenic pain disease patient have a rare NaV1.7 mutation?”. In this study, we have asked, “Do participants carrying monogenic pain disease mutations have an increased prevalence of chronic pain?”. The answer is no, at least for the *SCN9A* variants with more than 50 carriers in the UK Biobank cohort. These NaV1.7 mutations do not appear to be highly penetrant for pain. This contrasts with the view in the existing NaV1.7 literature and this seems primarily to be due to a degree of confirmation bias in the patient studies that have only sequenced the *SCN9A* gene and have only had partial view of the family pedigrees to track the genetic inheritance. Other potential causal variants and modulator genes have likely been missed as a consequence. The monogenic pain disease field would benefit from pooling data from consortia of electrophysiologists, clinicians, and geneticists to identify which of the *SCN9A* variants are truly causal for erythromelalgia and paroxysmal extreme pain disorder. A similar consortium effort has been undertaken for cystic fibrosis, where a curated database (https://cftr2.org) is maintained by an expert panel of clinicians, researchers, patient partners, and ethical advisors. It would be beneficial to establish a similar international monogenic pain disease cohort study/registry with whole-exome or genome sequencing. Sequencing of family members, regardless of disease status, would strengthen the evidence base for any genetic associations. In addition, patient DNA samples already banked could be whole exome or genome sequenced to re-evaluate the causal associations. Besides forming a useful resource for the patient, clinical and pain research community, it could also provide a point of access for investigators to recruit erythromelalgia patients for clinical trials – this is currently quite challenging due to their rarity and has likely contributed to the small number of therapeutic clinical trials for this disease group.

## Data Availability

The code to reproduce this analysis is available at https://github.com/graemenewton/Newton_2025_NaV_UKB

## Acknowledgements

This research has been conducted using data from UK Biobank, a major biomedical database under **project ID 64765**. The UK Biobank database received ethical approval from the North-West Haydock Research Ethics Committee (REC reference 21/NW/0157) and participants gave informed consent. We would like to thank all participants of the UK Biobank cohort who have provided necessary genetic and phenotypic information.

