## Supplementary Tables and Figures for "Carriers of *SCN9A* variants linked to inherited and acquired pain syndromes show no alteration in the prevalence of pain or analgesic usage in the UK Biobank cohort"

### Supplementary Material

| Source | Field ID | Question |
| --- | --- | --- |
| ORA | 3799_i0 | Headaches for 3+ months |
| ORA | 4067_i0 | Facial pains for 3+ months |
| ORA | 3404_i0 | Neck/shoulder pain for 3+ months |
| ORA | 3571_i0 | Back pain for 3+ months |
| ORA | 3741_i0 | Stomach/abdominal pain for 3+ months |
| ORA | 3414_i0 | Hip pain for 3+ months |
| ORA | 3773_i0 | Knee pain for 3+ months |
| ORA | 2956_i0 | General pain for 3+ months |

**Supplementary Table 1** Data fields used to generate the chronic pain phenotype group. ORA = Original Recruitment Assessment.

| Source | Field ID | Question |
| --- | --- | --- |
| EoPQ | 120046 | Burning pain present in pain that bothers the most |
| EoPQ | 120047 | Painful cold present in pain that bothers the most |
| EoPQ | 120048 | Electric shocks present in pain that bothers the most |
| EoPQ | 120049 | Tingling present in pain that bothers the most |
| EoPQ | 120050 | Pins and needles present in pain that bothers the most |
| EoPQ | 120051 | Numbness present in pain that bothers the most |
| EoPQ | 120052 | Itching present in pain that bothers the most |

**Supplementary Table 2** Data fields used to generate the neuropathic pain (DN4) phenotype group. EoPQ = Experience of Pain Questionnaire.

| Phenotype | Regular Expression Pattern |
| --- | --- |
| Gabapentinoid | "gabapentin\|pregabalin" |
| NaV_blocker | "mexiletine\|lacosamide\|carbamazepine\|oxcarbazepine\|  phenytoin\|sodium valproate\|lamotrigine" |
| Opioid_dual | "tramadol\|tapentadol" |
| Opioid_strong | "diamorphine\|morphine\|methadone\|fentanyl\|buprenorphine\|  oxycodone\|hydromorphone\|oramorph" |
| TCA | "amitriptyline\|nortriptyline\|protriptyline\|desipramine" |

**Supplementary Table 3** analgesic prescription phenotype groups and the regular expression patterns used to extract prescription records for each type of drug.

| Phenotype | Variable | Carriers | Carriers Pheno Rate | Controls | Controls Pheno Rate | Coef | SE | z | P | P FDR Adj. | OR | L95CI | U95CI |
| --- | --- | --- | --- | --- | --- | --- | --- | --- | --- | --- | --- | --- | --- |
| Chronic Pain | R185H | 1821 | 0.417 | 462158 | 0.418 | -0.017 | 0.048 | -0.355 | 0.723 | 0.948 | 0.983 | 0.895 | 1.080 |
| Chronic Pain | I228M | 1332 | 0.405 | 462158 | 0.418 | -0.033 | 0.056 | -0.582 | 0.560 | 0.948 | 0.968 | 0.867 | 1.080 |
| Chronic Pain | P610T | 24397 | 0.416 | 462158 | 0.418 | -0.001 | 0.013 | -0.101 | 0.920 | 0.948 | 0.999 | 0.973 | 1.025 |
| Chronic Pain | G616R | 74 | 0.554 | 462158 | 0.418 | 0.545 | 0.234 | 2.326 | 0.020 | 0.460 | 1.725 | 1.090 | 2.730 |
| Chronic Pain | K655R | 2415 | 0.406 | 462158 | 0.418 | -0.045 | 0.042 | -1.077 | 0.281 | 0.780 | 0.956 | 0.881 | 1.037 |
| Chronic Pain | W719C | 188 | 0.356 | 462158 | 0.418 | -0.416 | 0.154 | -2.703 | 0.007 | 0.460 | 0.660 | 0.488 | 0.892 |
| Chronic Pain | I720K | 244 | 0.381 | 462158 | 0.418 | -0.144 | 0.132 | -1.091 | 0.275 | 0.780 | 0.866 | 0.668 | 1.122 |
| Chronic Pain | I739V | 3716 | 0.424 | 462158 | 0.418 | 0.036 | 0.033 | 1.084 | 0.278 | 0.780 | 1.037 | 0.971 | 1.107 |
| Chronic Pain | V810M | 210 | 0.438 | 462158 | 0.418 | 0.080 | 0.140 | 0.570 | 0.568 | 0.948 | 1.083 | 0.823 | 1.425 |
| Chronic Pain | L1267V | 2282 | 0.422 | 462158 | 0.418 | 0.022 | 0.043 | 0.511 | 0.609 | 0.948 | 1.022 | 0.940 | 1.111 |
| Chronic Pain | W1538R | 392 | 0.413 | 462158 | 0.418 | -0.046 | 0.103 | -0.446 | 0.656 | 0.948 | 0.955 | 0.781 | 1.169 |
| Chronic Pain | (Intercept) |  |  |  |  | -0.599 | 0.021 | -28.560 | 0.000 |  | 0.549 | 0.527 | 0.572 |
| Chronic Pain | age |  |  |  |  | 0.006 | 0.000 | 17.371 | 0.000 |  | 1.006 | 1.006 | 1.007 |
| Chronic Pain | sex |  |  |  |  | -0.206 | 0.006 | -35.072 | 0.000 |  | 0.814 | 0.805 | 0.823 |
| Chronic Pain | PC1 |  |  |  |  | 0.001 | 0.000 | 10.403 | 0.000 |  | 1.001 | 1.000 | 1.001 |
| Chronic Pain | PC2 |  |  |  |  | 0.000 | 0.000 | -0.601 | 0.548 |  | 1.000 | 1.000 | 1.000 |
| Chronic Pain | PC3 |  |  |  |  | 0.003 | 0.000 | 12.811 | 0.000 |  | 1.003 | 1.002 | 1.003 |
| Chronic Pain | PC4 |  |  |  |  | 0.001 | 0.000 | 3.666 | 0.000 |  | 1.001 | 1.000 | 1.002 |
| Chronic Pain | PC5 |  |  |  |  | 0.002 | 0.000 | 5.622 | 0.000 |  | 1.002 | 1.001 | 1.003 |
| Chronic Pain | PC6 |  |  |  |  | 0.003 | 0.001 | 4.095 | 0.000 |  | 1.003 | 1.001 | 1.004 |
| Chronic Pain | PC7 |  |  |  |  | 0.004 | 0.001 | 6.062 | 0.000 |  | 1.004 | 1.002 | 1.005 |
| Chronic Pain | PC8 |  |  |  |  | 0.003 | 0.001 | 5.633 | 0.000 |  | 1.003 | 1.002 | 1.005 |
| Chronic Pain | PC9 |  |  |  |  | 0.001 | 0.001 | 1.955 | 0.051 |  | 1.001 | 1.000 | 1.003 |
| Chronic Pain | PC10 |  |  |  |  | 0.003 | 0.001 | 4.348 | 0.000 |  | 1.003 | 1.002 | 1.004 |
| Gabapentinoid | R185H | 575 | 0.104 | 148298 | 0.100 | 0.052 | 0.137 | 0.377 | 0.706 | 0.948 | 1.053 | 0.805 | 1.378 |
| Gabapentinoid | I228M | 445 | 0.076 | 148298 | 0.100 | -0.251 | 0.179 | -1.402 | 0.161 | 0.768 | 0.778 | 0.548 | 1.105 |
| Gabapentinoid | P610T | 7947 | 0.093 | 148298 | 0.100 | -0.081 | 0.040 | -2.041 | 0.041 | 0.460 | 0.922 | 0.853 | 0.997 |
| Gabapentinoid | G616R | 29 | 0.172 | 148298 | 0.100 | 0.607 | 0.493 | 1.231 | 0.218 | 0.780 | 1.835 | 0.698 | 4.826 |
| Gabapentinoid | K655R | 804 | 0.100 | 148298 | 0.100 | -0.006 | 0.118 | -0.049 | 0.961 | 0.973 | 0.994 | 0.788 | 1.254 |
| Gabapentinoid | W719C | 45 | 0.067 | 148298 | 0.100 | -0.543 | 0.601 | -0.903 | 0.367 | 0.837 | 0.581 | 0.179 | 1.888 |
| Gabapentinoid | I720K | 78 | 0.103 | 148298 | 0.100 | 0.051 | 0.374 | 0.138 | 0.891 | 0.948 | 1.053 | 0.506 | 2.191 |
| Gabapentinoid | I739V | 1293 | 0.104 | 148298 | 0.100 | 0.068 | 0.092 | 0.745 | 0.456 | 0.878 | 1.071 | 0.895 | 1.281 |
| Gabapentinoid | V810M | 63 | 0.095 | 148298 | 0.100 | -0.054 | 0.430 | -0.126 | 0.900 | 0.948 | 0.947 | 0.407 | 2.202 |
| Gabapentinoid | L1267V | 809 | 0.096 | 148298 | 0.100 | -0.028 | 0.120 | -0.236 | 0.814 | 0.948 | 0.972 | 0.769 | 1.229 |
| Gabapentinoid | W1538R | 135 | 0.156 | 148298 | 0.100 | 0.458 | 0.238 | 1.921 | 0.055 | 0.527 | 1.580 | 0.991 | 2.521 |
| Gabapentinoid | (Intercept) |  |  |  |  | -2.752 | 0.064 | -43.107 | 0.000 |  | 0.064 | 0.056 | 0.072 |
| Gabapentinoid | age |  |  |  |  | 0.012 | 0.001 | 10.564 | 0.000 |  | 1.012 | 1.009 | 1.014 |
| Gabapentinoid | sex |  |  |  |  | -0.298 | 0.017 | -17.209 | 0.000 |  | 0.742 | 0.717 | 0.768 |
| Gabapentinoid | PC1 |  |  |  |  | 0.000 | 0.000 | 1.918 | 0.055 |  | 1.000 | 1.000 | 1.001 |
| Gabapentinoid | PC2 |  |  |  |  | 0.001 | 0.000 | 1.604 | 0.109 |  | 1.001 | 1.000 | 1.001 |
| Gabapentinoid | PC3 |  |  |  |  | 0.005 | 0.001 | 6.578 | 0.000 |  | 1.005 | 1.003 | 1.006 |
| Gabapentinoid | PC4 |  |  |  |  | 0.000 | 0.001 | 0.343 | 0.731 |  | 1.000 | 0.999 | 1.002 |
| Gabapentinoid | PC5 |  |  |  |  | 0.008 | 0.001 | 7.159 | 0.000 |  | 1.008 | 1.005 | 1.010 |
| Gabapentinoid | PC6 |  |  |  |  | -0.003 | 0.002 | -1.242 | 0.214 |  | 0.997 | 0.993 | 1.002 |
| Gabapentinoid | PC7 |  |  |  |  | -0.001 | 0.002 | -0.533 | 0.594 |  | 0.999 | 0.996 | 1.002 |
| Gabapentinoid | PC8 |  |  |  |  | 0.002 | 0.002 | 1.117 | 0.264 |  | 1.002 | 0.998 | 1.006 |
| Gabapentinoid | PC9 |  |  |  |  | 0.010 | 0.002 | 4.071 | 0.000 |  | 1.010 | 1.005 | 1.015 |
| Gabapentinoid | PC10 |  |  |  |  | 0.007 | 0.002 | 3.546 | 0.000 |  | 1.007 | 1.003 | 1.011 |
| NaV Blocker | R185H | 575 | 0.026 | 148298 | 0.031 | -0.213 | 0.271 | -0.787 | 0.431 | 0.874 | 0.808 | 0.475 | 1.375 |
| NaV Blocker | I228M | 445 | 0.031 | 148298 | 0.031 | 0.038 | 0.272 | 0.140 | 0.889 | 0.948 | 1.039 | 0.609 | 1.771 |
| NaV Blocker | P610T | 7947 | 0.031 | 148298 | 0.031 | 0.006 | 0.067 | 0.096 | 0.924 | 0.948 | 1.006 | 0.883 | 1.147 |
| NaV Blocker | G616R | 29 | 0.000 | 148298 | 0.031 | -10.121 | 99.333 | -0.102 | 0.919 | 0.948 | 0.000 | 0.000 | Inf |
| NaV Blocker | K655R | 804 | 0.039 | 148298 | 0.031 | 0.234 | 0.184 | 1.272 | 0.203 | 0.780 | 1.264 | 0.881 | 1.812 |
| NaV Blocker | W719C | 45 | 0.000 | 148298 | 0.031 | -9.925 | 79.731 | -0.124 | 0.901 | 0.948 | 0.000 | 0.000 | Inf |
| NaV Blocker | I720K | 78 | 0.051 | 148298 | 0.031 | 0.546 | 0.514 | 1.063 | 0.288 | 0.780 | 1.727 | 0.631 | 4.727 |
| NaV Blocker | I739V | 1293 | 0.022 | 148298 | 0.031 | -0.351 | 0.192 | -1.829 | 0.067 | 0.576 | 0.704 | 0.484 | 1.025 |
| NaV Blocker | V810M | 63 | 0.048 | 148298 | 0.031 | 0.489 | 0.592 | 0.825 | 0.409 | 0.851 | 1.630 | 0.511 | 5.204 |
| NaV Blocker | L1267V | 809 | 0.022 | 148298 | 0.031 | -0.328 | 0.239 | -1.373 | 0.170 | 0.768 | 0.720 | 0.451 | 1.150 |
| NaV Blocker | W1538R | 135 | 0.037 | 148298 | 0.031 | 0.204 | 0.456 | 0.448 | 0.654 | 0.948 | 1.227 | 0.502 | 3.000 |
| NaV Blocker | (Intercept) |  |  |  |  | -3.663 | 0.110 | -33.326 | 0.000 |  | 0.026 | 0.021 | 0.032 |
| NaV Blocker | age |  |  |  |  | 0.005 | 0.002 | 2.400 | 0.016 |  | 1.005 | 1.001 | 1.008 |
| NaV Blocker | sex |  |  |  |  | -0.165 | 0.030 | -5.520 | 0.000 |  | 0.848 | 0.799 | 0.899 |
| NaV Blocker | PC1 |  |  |  |  | -0.001 | 0.000 | -2.122 | 0.034 |  | 0.999 | 0.999 | 1.000 |
| NaV Blocker | PC2 |  |  |  |  | 0.001 | 0.001 | 1.392 | 0.164 |  | 1.001 | 1.000 | 1.002 |
| NaV Blocker | PC3 |  |  |  |  | 0.000 | 0.001 | -0.197 | 0.844 |  | 1.000 | 0.997 | 1.002 |
| NaV Blocker | PC4 |  |  |  |  | 0.000 | 0.001 | -0.199 | 0.842 |  | 1.000 | 0.997 | 1.003 |
| NaV Blocker | PC5 |  |  |  |  | 0.002 | 0.002 | 1.065 | 0.287 |  | 1.002 | 0.998 | 1.006 |
| NaV Blocker | PC6 |  |  |  |  | -0.004 | 0.004 | -1.032 | 0.302 |  | 0.996 | 0.989 | 1.003 |
| NaV Blocker | PC7 |  |  |  |  | -0.004 | 0.003 | -1.360 | 0.174 |  | 0.996 | 0.989 | 1.002 |
| NaV Blocker | PC8 |  |  |  |  | -0.005 | 0.003 | -1.519 | 0.129 |  | 0.995 | 0.988 | 1.002 |
| NaV Blocker | PC9 |  |  |  |  | 0.007 | 0.004 | 1.602 | 0.109 |  | 1.007 | 0.998 | 1.015 |
| NaV Blocker | PC10 |  |  |  |  | 0.003 | 0.004 | 0.698 | 0.485 |  | 1.003 | 0.995 | 1.010 |
| Neuropathic Pain | R185H | 271 | 0.170 | 69790 | 0.181 | -0.080 | 0.162 | -0.493 | 0.622 | 0.948 | 0.923 | 0.672 | 1.269 |
| Neuropathic Pain | I228M | 202 | 0.183 | 69790 | 0.181 | 0.041 | 0.182 | 0.223 | 0.823 | 0.948 | 1.041 | 0.729 | 1.489 |
| Neuropathic Pain | P610T | 3848 | 0.173 | 69790 | 0.181 | -0.042 | 0.044 | -0.954 | 0.340 | 0.837 | 0.959 | 0.880 | 1.045 |
| Neuropathic Pain | G616R | 12 | 0.083 | 69790 | 0.181 | -0.869 | 1.044 | -0.832 | 0.405 | 0.851 | 0.419 | 0.054 | 3.246 |
| Neuropathic Pain | K655R | 409 | 0.191 | 69790 | 0.181 | 0.062 | 0.126 | 0.493 | 0.622 | 0.948 | 1.064 | 0.831 | 1.363 |
| Neuropathic Pain | W719C | 10 | 0.200 | 69790 | 0.181 | -0.212 | 0.797 | -0.265 | 0.791 | 0.948 | 0.809 | 0.170 | 3.861 |
| Neuropathic Pain | I720K | 45 | 0.133 | 69790 | 0.181 | -0.334 | 0.439 | -0.760 | 0.447 | 0.878 | 0.716 | 0.303 | 1.693 |
| Neuropathic Pain | I739V | 608 | 0.173 | 69790 | 0.181 | -0.043 | 0.108 | -0.397 | 0.691 | 0.948 | 0.958 | 0.776 | 1.184 |
| Neuropathic Pain | V810M | 41 | 0.195 | 69790 | 0.181 | 0.098 | 0.395 | 0.247 | 0.805 | 0.948 | 1.103 | 0.509 | 2.390 |
| Neuropathic Pain | L1267V | 338 | 0.160 | 69790 | 0.181 | -0.143 | 0.149 | -0.964 | 0.335 | 0.837 | 0.866 | 0.647 | 1.160 |
| Neuropathic Pain | W1538R | 70 | 0.286 | 69790 | 0.181 | 0.571 | 0.265 | 2.153 | 0.031 | 0.460 | 1.770 | 1.053 | 2.976 |
| Neuropathic Pain | (Intercept) |  |  |  |  | -1.247 | 0.071 | -17.612 | 0.000 |  | 0.287 | 0.250 | 0.330 |
| Neuropathic Pain | age |  |  |  |  | -0.004 | 0.001 | -3.436 | 0.001 |  | 0.996 | 0.993 | 0.998 |
| Neuropathic Pain | sex |  |  |  |  | -0.014 | 0.019 | -0.714 | 0.475 |  | 0.986 | 0.949 | 1.025 |
| Neuropathic Pain | PC1 |  |  |  |  | 0.001 | 0.000 | 5.218 | 0.000 |  | 1.001 | 1.001 | 1.002 |
| Neuropathic Pain | PC2 |  |  |  |  | -0.001 | 0.000 | -2.098 | 0.036 |  | 0.999 | 0.998 | 1.000 |
| Neuropathic Pain | PC3 |  |  |  |  | 0.001 | 0.001 | 1.137 | 0.256 |  | 1.001 | 0.999 | 1.003 |
| Neuropathic Pain | PC4 |  |  |  |  | 0.002 | 0.001 | 2.032 | 0.042 |  | 1.002 | 1.000 | 1.004 |
| Neuropathic Pain | PC5 |  |  |  |  | 0.007 | 0.001 | 5.075 | 0.000 |  | 1.007 | 1.004 | 1.009 |
| Neuropathic Pain | PC6 |  |  |  |  | 0.001 | 0.003 | 0.503 | 0.615 |  | 1.001 | 0.996 | 1.007 |
| Neuropathic Pain | PC7 |  |  |  |  | 0.005 | 0.002 | 2.466 | 0.014 |  | 1.005 | 1.001 | 1.009 |
| Neuropathic Pain | PC8 |  |  |  |  | 0.007 | 0.002 | 2.817 | 0.005 |  | 1.007 | 1.002 | 1.011 |
| Neuropathic Pain | PC9 |  |  |  |  | -0.003 | 0.002 | -1.585 | 0.113 |  | 0.997 | 0.992 | 1.001 |
| Neuropathic Pain | PC10 |  |  |  |  | -0.002 | 0.003 | -0.741 | 0.458 |  | 0.998 | 0.992 | 1.004 |
| Opioid (dual) | R185H | 575 | 0.191 | 148298 | 0.180 | 0.075 | 0.107 | 0.702 | 0.483 | 0.906 | 1.078 | 0.874 | 1.328 |
| Opioid (dual) | I228M | 445 | 0.153 | 148298 | 0.180 | -0.163 | 0.132 | -1.234 | 0.217 | 0.780 | 0.849 | 0.655 | 1.101 |
| Opioid (dual) | P610T | 7947 | 0.173 | 148298 | 0.180 | -0.043 | 0.031 | -1.404 | 0.160 | 0.768 | 0.958 | 0.902 | 1.017 |
| Opioid (dual) | G616R | 29 | 0.345 | 148298 | 0.180 | 0.858 | 0.393 | 2.185 | 0.029 | 0.460 | 2.359 | 1.093 | 5.094 |
| Opioid (dual) | K655R | 804 | 0.180 | 148298 | 0.180 | -0.002 | 0.092 | -0.016 | 0.987 | 0.987 | 0.998 | 0.833 | 1.196 |
| Opioid (dual) | W719C | 45 | 0.133 | 148298 | 0.180 | -0.501 | 0.442 | -1.134 | 0.257 | 0.780 | 0.606 | 0.255 | 1.441 |
| Opioid (dual) | I720K | 78 | 0.218 | 148298 | 0.180 | 0.253 | 0.275 | 0.918 | 0.359 | 0.837 | 1.288 | 0.751 | 2.208 |
| Opioid (dual) | I739V | 1293 | 0.196 | 148298 | 0.180 | 0.120 | 0.071 | 1.708 | 0.088 | 0.675 | 1.128 | 0.982 | 1.295 |
| Opioid (dual) | V810M | 63 | 0.238 | 148298 | 0.180 | 0.351 | 0.297 | 1.183 | 0.237 | 0.780 | 1.421 | 0.794 | 2.543 |
| Opioid (dual) | L1267V | 809 | 0.195 | 148298 | 0.180 | 0.109 | 0.089 | 1.221 | 0.222 | 0.780 | 1.115 | 0.936 | 1.328 |
| Opioid (dual) | W1538R | 135 | 0.156 | 148298 | 0.180 | -0.209 | 0.238 | -0.877 | 0.380 | 0.837 | 0.812 | 0.509 | 1.294 |
| Opioid (dual) | (Intercept) |  |  |  |  | -2.484 | 0.050 | -49.419 | 0.000 |  | 0.083 | 0.076 | 0.092 |
| Opioid (dual) | age |  |  |  |  | 0.018 | 0.001 | 21.248 | 0.000 |  | 1.018 | 1.017 | 1.020 |
| Opioid (dual) | sex |  |  |  |  | -0.212 | 0.013 | -15.832 | 0.000 |  | 0.809 | 0.788 | 0.830 |
| Opioid (dual) | PC1 |  |  |  |  | 0.001 | 0.000 | 3.768 | 0.000 |  | 1.001 | 1.000 | 1.001 |
| Opioid (dual) | PC2 |  |  |  |  | 0.001 | 0.000 | 2.719 | 0.007 |  | 1.001 | 1.000 | 1.001 |
| Opioid (dual) | PC3 |  |  |  |  | 0.003 | 0.001 | 5.859 | 0.000 |  | 1.003 | 1.002 | 1.004 |
| Opioid (dual) | PC4 |  |  |  |  | 0.001 | 0.001 | 2.103 | 0.036 |  | 1.001 | 1.000 | 1.003 |
| Opioid (dual) | PC5 |  |  |  |  | 0.005 | 0.001 | 5.800 | 0.000 |  | 1.005 | 1.003 | 1.006 |
| Opioid (dual) | PC6 |  |  |  |  | 0.000 | 0.002 | -0.102 | 0.919 |  | 1.000 | 0.997 | 1.003 |
| Opioid (dual) | PC7 |  |  |  |  | 0.001 | 0.001 | 0.635 | 0.526 |  | 1.001 | 0.998 | 1.004 |
| Opioid (dual) | PC8 |  |  |  |  | 0.005 | 0.001 | 3.700 | 0.000 |  | 1.005 | 1.003 | 1.008 |
| Opioid (dual) | PC9 |  |  |  |  | 0.007 | 0.002 | 3.693 | 0.000 |  | 1.007 | 1.003 | 1.011 |
| Opioid (dual) | PC10 |  |  |  |  | 0.005 | 0.002 | 3.343 | 0.001 |  | 1.005 | 1.002 | 1.008 |
| Opioid (strong) | R185H | 575 | 0.043 | 148298 | 0.055 | -0.229 | 0.205 | -1.114 | 0.265 | 0.780 | 0.796 | 0.532 | 1.190 |
| Opioid (strong) | I228M | 445 | 0.056 | 148298 | 0.055 | 0.026 | 0.207 | 0.126 | 0.900 | 0.948 | 1.026 | 0.684 | 1.539 |
| Opioid (strong) | P610T | 7947 | 0.051 | 148298 | 0.055 | -0.075 | 0.052 | -1.435 | 0.151 | 0.768 | 0.928 | 0.837 | 1.028 |
| Opioid (strong) | G616R | 29 | 0.069 | 148298 | 0.055 | 0.233 | 0.735 | 0.317 | 0.751 | 0.948 | 1.263 | 0.299 | 5.337 |
| Opioid (strong) | K655R | 804 | 0.053 | 148298 | 0.055 | -0.045 | 0.158 | -0.286 | 0.775 | 0.948 | 0.956 | 0.702 | 1.302 |
| Opioid (strong) | W719C | 45 | 0.044 | 148298 | 0.055 | -0.186 | 0.730 | -0.255 | 0.799 | 0.948 | 0.830 | 0.198 | 3.472 |
| Opioid (strong) | I720K | 78 | 0.051 | 148298 | 0.055 | -0.083 | 0.515 | -0.161 | 0.872 | 0.948 | 0.920 | 0.336 | 2.524 |
| Opioid (strong) | I739V | 1293 | 0.069 | 148298 | 0.055 | 0.226 | 0.111 | 2.036 | 0.042 | 0.460 | 1.253 | 1.008 | 1.557 |
| Opioid (strong) | V810M | 63 | 0.048 | 148298 | 0.055 | -0.186 | 0.593 | -0.314 | 0.754 | 0.948 | 0.830 | 0.260 | 2.653 |
| Opioid (strong) | L1267V | 809 | 0.058 | 148298 | 0.055 | 0.049 | 0.151 | 0.321 | 0.748 | 0.948 | 1.050 | 0.781 | 1.412 |
| Opioid (strong) | W1538R | 135 | 0.037 | 148298 | 0.055 | -0.406 | 0.457 | -0.889 | 0.374 | 0.837 | 0.666 | 0.272 | 1.631 |
| Opioid (strong) | (Intercept) |  |  |  |  | -5.113 | 0.092 | -55.853 | 0.000 |  | 0.006 | 0.005 | 0.007 |
| Opioid (strong) | age |  |  |  |  | 0.039 | 0.002 | 25.449 | 0.000 |  | 1.040 | 1.037 | 1.043 |
| Opioid (strong) | sex |  |  |  |  | -0.028 | 0.022 | -1.273 | 0.203 |  | 0.972 | 0.930 | 1.015 |
| Opioid (strong) | PC1 |  |  |  |  | 0.000 | 0.000 | -0.833 | 0.405 |  | 1.000 | 0.999 | 1.000 |
| Opioid (strong) | PC2 |  |  |  |  | 0.003 | 0.001 | 4.487 | 0.000 |  | 1.003 | 1.002 | 1.004 |
| Opioid (strong) | PC3 |  |  |  |  | 0.001 | 0.001 | 0.509 | 0.610 |  | 1.001 | 0.998 | 1.003 |
| Opioid (strong) | PC4 |  |  |  |  | 0.001 | 0.001 | 0.551 | 0.581 |  | 1.001 | 0.998 | 1.003 |
| Opioid (strong) | PC5 |  |  |  |  | -0.001 | 0.001 | -1.008 | 0.313 |  | 0.999 | 0.996 | 1.001 |
| Opioid (strong) | PC6 |  |  |  |  | -0.003 | 0.004 | -0.821 | 0.412 |  | 0.997 | 0.990 | 1.004 |
| Opioid (strong) | PC7 |  |  |  |  | 0.001 | 0.002 | 0.571 | 0.568 |  | 1.001 | 0.997 | 1.006 |
| Opioid (strong) | PC8 |  |  |  |  | 0.004 | 0.003 | 1.265 | 0.206 |  | 1.004 | 0.998 | 1.009 |
| Opioid (strong) | PC9 |  |  |  |  | 0.004 | 0.003 | 1.247 | 0.213 |  | 1.004 | 0.998 | 1.010 |
| Opioid (strong) | PC10 |  |  |  |  | 0.009 | 0.003 | 2.827 | 0.005 |  | 1.009 | 1.003 | 1.015 |
| TCA | R185H | 575 | 0.263 | 148298 | 0.260 | 0.015 | 0.096 | 0.153 | 0.878 | 0.948 | 1.015 | 0.841 | 1.225 |
| TCA | I228M | 445 | 0.249 | 148298 | 0.260 | -0.033 | 0.111 | -0.299 | 0.765 | 0.948 | 0.967 | 0.779 | 1.202 |
| TCA | P610T | 7947 | 0.254 | 148298 | 0.260 | -0.028 | 0.027 | -1.050 | 0.294 | 0.780 | 0.972 | 0.923 | 1.025 |
| TCA | G616R | 29 | 0.310 | 148298 | 0.260 | 0.178 | 0.406 | 0.439 | 0.661 | 0.948 | 1.195 | 0.539 | 2.647 |
| TCA | K655R | 804 | 0.256 | 148298 | 0.260 | -0.018 | 0.082 | -0.221 | 0.825 | 0.948 | 0.982 | 0.837 | 1.153 |
| TCA | W719C | 45 | 0.178 | 148298 | 0.260 | -0.582 | 0.394 | -1.475 | 0.140 | 0.768 | 0.559 | 0.258 | 1.211 |
| TCA | I720K | 78 | 0.192 | 148298 | 0.260 | -0.423 | 0.289 | -1.461 | 0.144 | 0.768 | 0.655 | 0.372 | 1.155 |
| TCA | I739V | 1293 | 0.286 | 148298 | 0.260 | 0.137 | 0.063 | 2.194 | 0.028 | 0.460 | 1.147 | 1.015 | 1.297 |
| TCA | V810M | 63 | 0.238 | 148298 | 0.260 | -0.086 | 0.299 | -0.289 | 0.772 | 0.948 | 0.917 | 0.511 | 1.647 |
| TCA | L1267V | 809 | 0.282 | 148298 | 0.260 | 0.119 | 0.079 | 1.498 | 0.134 | 0.768 | 1.126 | 0.964 | 1.315 |
| TCA | W1538R | 135 | 0.237 | 148298 | 0.260 | -0.139 | 0.204 | -0.679 | 0.497 | 0.912 | 0.871 | 0.583 | 1.299 |
| TCA | (Intercept) |  |  |  |  | -1.240 | 0.043 | -28.668 | 0.000 |  | 0.289 | 0.266 | 0.315 |
| TCA | age |  |  |  |  | 0.008 | 0.001 | 10.223 | 0.000 |  | 1.008 | 1.006 | 1.009 |
| TCA | sex |  |  |  |  | -0.626 | 0.012 | -51.984 | 0.000 |  | 0.535 | 0.522 | 0.548 |
| TCA | PC1 |  |  |  |  | 0.000 | 0.000 | 0.519 | 0.604 |  | 1.000 | 1.000 | 1.000 |
| TCA | PC2 |  |  |  |  | 0.001 | 0.000 | 3.826 | 0.000 |  | 1.001 | 1.000 | 1.002 |
| TCA | PC3 |  |  |  |  | 0.004 | 0.000 | 8.127 | 0.000 |  | 1.004 | 1.003 | 1.005 |
| TCA | PC4 |  |  |  |  | 0.000 | 0.001 | -0.569 | 0.570 |  | 1.000 | 0.999 | 1.001 |
| TCA | PC5 |  |  |  |  | -0.003 | 0.001 | -4.208 | 0.000 |  | 0.997 | 0.995 | 0.998 |
| TCA | PC6 |  |  |  |  | -0.001 | 0.001 | -0.473 | 0.636 |  | 0.999 | 0.997 | 1.002 |
| TCA | PC7 |  |  |  |  | 0.001 | 0.001 | 0.867 | 0.386 |  | 1.001 | 0.999 | 1.003 |
| TCA | PC8 |  |  |  |  | 0.002 | 0.001 | 1.693 | 0.090 |  | 1.002 | 1.000 | 1.005 |
| TCA | PC9 |  |  |  |  | 0.009 | 0.002 | 5.567 | 0.000 |  | 1.009 | 1.006 | 1.013 |
| TCA | PC10 |  |  |  |  | 0.005 | 0.001 | 3.606 | 0.000 |  | 1.005 | 1.002 | 1.008 |

**Supplementary Table 4** – Logistic regression model summaries for the effects of Na_V_1.7 mutations R185H, I228M, P610T, G616R, K655R, W719C, I720K, I739V, V810M, L1267V, and W1538R on the risk of chronic pain, neuropathic pain, gabapentinoid, Na_V_ blockers, dual mechanism opioids, strong opioids, and tricyclic antidepressants.

| Na_V_1.7 Mutation | Variant ID | Ephys GoF | UKB | Disease | Paralogue GoF Ephys | Diseases associated with Paralogues | Refs |
| --- | --- | --- | --- | --- | --- | --- | --- |
| Q10K | - | N | N | EM |  |  | ^82^ |
| I136V | 2:166306571:T:C | Y | N | EM | SCN4 | **SCN**1A: Dravets: **SCN**4A: Paramyotonia congenita; **SCN**5A: Ventricular arrhythmia | ^83,84^ |
| S211P |  | Y | N | EM |  | **SCN**2A: Malignant migrating partial seizures of infancy; | ^85^ |
| F216S | 2:166304279:A:G | Y | N | EM |  | **SCN**1A: neurodevelopmental disorder, Generalized epilepsy with febrile seizures | ^68,86^ |
| I234T | 2:166303290:A:G | Y | N | EM |  | **SCN**2A: Encephalopathy with early-onset epilepsy; **SCN**8A:Epileptic encephalopathy; **SCN**5A:Long QT syndrome | ^87^ |
| S241T | 2:166303270:A:T | Y | N | EM | SCN10 | **SCN**1A: Dravet syndrome | ^88,89^ |
| L245V |  | Y | N | EM |  | **SCN**2A:PEHO syndrome; **SCN**3A:Epilepsy- focal; **SCN**8A:Epileptic encephalopathy- early infantile; **SCN**4A: Isolated eyelid closure myotonia; **SCN**5A:Brugada syndrome | ^19^ |
| N395K | 2:166288566:G:A, 2:166288566:G:C | N | N | EM | SCN4 | **SCN**1A:Early infantile epileptic encephalopathy; **SCN**4A:Hyperkalemic Periodic Paralysis Type 1,Myotonia; **SCN**5A:Congenital long QT syndrome, Brugada syndrome | ^68^ |
| V400M | 2:166288553:C:T | Y | N | EM | SCN2, 4 & 5 | **SCN**1A: Dravet syndrome; **SCN**2A:Early infantile epileptic encephalopathy **SCN**4A:Congenital myasthenic syndrome;Hyperkalemic Periodic Paralysis Type 1;Paramyotonia congenita; **SCN**5A:Brugada syndrome; Congenital long QT syndrome; | ^90^ |
| D623N | 2:166284560:G:A | Y | N | SFN |  | **SCN**2A:Dravet syndrome | ^58^ |
| L823R |  | Y | N | EM |  | **SCN**1A:Dravet syndrome; | ^91,92^ |
| F826Y |  | Y | N | EM |  | **SCN**8A:Early infantile epileptic encephalopathy; **SCN**4A:Myotonia- non-dystrophic | ^93^ |
| I848T | 2:166277281:A:G | Y | N | EM | SCN3 | **SCN**3A: Early infantile epileptic encephalopathy; **SCN**8A:Early infantile epileptic encephalopathy ; **SCN**4A:Myotonia,Hyperkalemic Periodic Paralysis Type 1;Paramyotonia congenita of von Eulenburg; | ^8,94^ |
| G856R | 2:166277258:C:G | Y | N | EM |  | **SCN**2A:Encephalopathy with early-onset epilepsy; **SCN**5A:Long QT syndrome | ^95^ |
| G856D | 2:166277257:C:T | Y | N | EM |  |  | ^96^ |
| L858H | 2:166277251:A:T | Y | N | EM |  | **SCN**2A:Neurodevelopmental disorder- epilepsy-related; | ^8,97^ |
| L858F | 2:166277252:G:A | Y | N | EM |  |  | ^68,98^ |
| A863P | 2:166277237:G:C | Y | N | EM |  |  | ^99^ |
| V872G |  | Y | N | EM |  | **SCN**1A:Focal epilepsy; **SCN**8A:Early infantile epileptic encephalopathy; **SCN**4A:Myotonia- non-dystrophic | ^100^ |
| Q875E |  | Y | N | EM |  | **SCN**1A:Dravet syndrome; **SCN**2A: Neurodevelopmental disorder- epilepsy-related | ^27,101^ |
| L955del |  | Y | N | EM |  | **SCN**1A:Dravet syndrome | ^37^ |
| V1298D | 2:166228971:A:T | N | N | PEPD |  | **SCN**1A:Severe myoclonic epilepsy of infancy; **SCN**2A:Episodic ataxia; **SCN**8A: Epileptic encephalopathy- infantile | ^13^ |
| V1298F | 2:166228972:C:A, 2:166228972:C:T | Y | N | PEPD |  |  | ^13,102^ |
| V1299F | 2:166228969:C:T | Y | N | PEPD |  | **SCN**2A:Ohtahara syndrome, Migrating focal seizures of infancy; **SCN**4A:Episodic paralyses and myotonic discharges; **SCN**5A:Brugada syndrome | ^13,102^ |
| P1308L |  | Y | N | EM | SCN3, 4 & 5 | **SCN**1A:Epileptic encephalopathy- early onset; **SCN**3A:Early infantile epileptic encephalopathy **SCN**4A:Myotonia,Hypokalemic periodic paralysis; **SCN**5A:Long QT syndrome | ^72^ |
| V1316A |  | Y | N | EM | SCN11 | **SCN**1A:Generalized epilepsy with febrile seizures plus; **SCN**4A:Myotonia; **SCN**5A: Brugada syndrome; **SCN**11A:Cold-aggravated peripheral pain | ^94,103^ |
| F1449V |  | Y | N | EM | SCN4 & 5 | **SCN**1A:Epileptic encephalopathy- early infantile; **SCN**5A:Long QT syndrome | ^64^ |
| I1461T | 2:166204448:A:G, 2:166204448:A:T | Y | N | PEPD | SCN1 (LoF), SCN4 (LoF) | **SCN**1A:Epilepsy and/or neurodevelopmental disorders; **SCN**2A: Neurodevelopmental disorder- epilepsy-related; **SCN**8A:Early infantile epileptic encephalopathy; **SCN**5A:Brugada syndrome | ^13,102^ |
| F1462V | 2:166204446:A:C | Y | N | PEPD |  | **SCN**1A:Hemiplegic migraine ; **SCN**5A:Cardiac conduction disorder, Sudden adult death syndrome | ^13^ |
| T1464I | 2:166204439:G:A | Y | N | PEPD | SCN3 & 4 | **SCN**4A Congenital myasthenic syndrome; Hyperkalemic Periodic Paralysis Type 1;Paramyotonia congenita of von Eulenburg; **SCN**5A:Long QT syndrome | ^13^ |
| G1607R |  | Y | N | PEPD | SCN5 | **SCN**1A:Epilepsy and/or neurodevelopmental disorders; **SCN**2A:Ohtahara syndrome,Episodic ataxia;**SCN**8A:Early infantile epileptic encephalopathy; **SCN**4A:Paramyotonia congenita of von Eulenburg; **SCN**5A:Long QT syndrome- malignant perinatal variant | ^104^ |
| L1612P | 2:166199771:A:G | Y | N | PEPD |  | **SCN**1A:Hemiplegic migraine; **SCN**8A:Seizures; **SCN**4A:Myotonia | ^105^ |
| F1624S | 2:166199735:A:G | Y | N | PEPD |  | **SCN**1A:Severe myoclonic epilepsy of infancy,Hemiplegic migraine; **SCN**2A:Encephalopathy with early-onset epilepsy; **SCN**4A:Paramyotonia congenita | ^69^ |
| M1627K |  | Y | N | PEPD |  | **SCN**1A:Dravet syndrome; **SCN**8A:Partial seizures with intellectual / developmental disabilities; **SCN**4A:Paramyotonia congenita of von Eulenburg, | ^106^ |
| A1632E | 2:166199711:C:A | Y | N | EM | SCN5 | **SCN**1A:Partial seizures of infancy- malignant migrating, **SCN**2A:Benign familial neonatal-infantile seizures;Early infantile epileptic encephalopathy;**SCN**8A:Early infantile epileptic encephalopathy; **SCN**4A:Hyperkalemic Periodic Paralysis Type 1,Myotonia | ^107^ |
| A1632T | 2:166199711:C:T | Y | N | EM |  |  | ^20^ |
| A1632G |  | Y | N | EM | SCN5 | **SCN**1A:Partial seizures of infancy- malignant migrating, **SCN**2A:Benign familial neonatal-infantile seizures;Early infantile epileptic encephalopathy;**SCN**8A:Early infantile epileptic encephalopathy; **SCN**4A:Hyperkalemic Periodic Paralysis Type 1,Myotonia | ^108^ |
| V1740L | 2:166199388:C:G | N | N | PEPD | SCN4 | **SCN**8A:**SCN**8A-related disorder;**SCN**4A:Hyperkalemic Periodic Paralysis Type 1;Paramyotonia congenita of von Eulenburg;,Myotonia;**SCN**5A:Long QT syndrome | ^109^ |
| A1746G |  | Y | N | EM |  | **SCN**1A:Severe myoclonic epilepsy of infancy;**SCN**2A:Benign familial neonatal-infantile seizures;Early infantile epileptic encephalopathy | ^10^ |
| In UKB | | | | | | | |
| Q10R | 2:166311728:T:C | Y | Y | EM |  |  | ^9^ |
| R185H | 2:166305834:C:T | Y | Y | SFN |  | **SCN**2A:Benign familial neonatal-infantile seizures,Febrile and afebrile seizures; **SCN**5A:Long QT syndrome | ^60,58^ |
| P187L | 2:166305828:G:A | N | Y | EM |  |  | ^53^ |
| I228M | 2:166304242:G:C | Y | Y | EM |  |  | ^52^ |
| P610T | 2:166284599:G:T | Y | Y | EM* |  |  | ^110,111^ |
| G616R | 2:166284581:C:T | Y | Y | EM |  |  | ^62^ |
| N641Y | 2:166284506:T:A | Y | Y | EP |  |  | ^33^ |
| K655R | 2:166281786:T:C | Y | Y | EP |  | **SCN**5A:Long QT syndrome | ^33^ |
| W719C | 2:166280510:C:G, 2:166280510:C:A | Y | Y | SFN |  | **SCN**1A:Dravet syndrome | ^112^ |
| I720K | 2:166280508:A:T | Y | Y | SFN |  |  | ^58^ |
| I739V | 2:166280452:T:C | Y | Y | SFN |  | **SCN**10A:Brugada syndrome | ^112^ |
| V810M | 2:166278196:C:T | Y | Y | SFN |  |  | ^77^ |
| M932L | 2:166277030:T:G | Y | Y | SFN |  | **SCN**2A:Epilepsy- benign neonatal-infantile | ^58^ |
| V991L | 2:166272746:C:A | Y | Y | EM |  |  | ^54^ |
| R996C | 2:166272731:G:A | N | Y | PEPD |  |  | ^22^ |
| R1150W | 2:166242648:A:G | Y | Y | EM* |  |  | ^113,114^ |
| L1267V | 2:166233432:G:C | Y | Y | SFN |  |  | ^112^ |
| M1532I | 2:166204100:C:T | Y | Y | SFN |  |  | ^58^ |
| W1538R | 2:166204084:A:G | Y | Y | EM |  | **SCN**1A:Rasmussen encephalitis | ^10^ |

**Supplementary Table 5** List of Na_V_1.7 gain-of-function channelopathy mutations, their disease association, sorted by their presence within UK Biobank, and whether the mutation has had an electrophysiological gain-of-function confirmed. Highlighted variants had either ≥50 carriers or were excluded because there were too many carriers. The table contains all published erythromelalgia and PEPD mutations. A selection of small fibre neuropathy mutations with gain-of-function evidence is also presented. If functional evidence was not published at the time of discovery of the mutation, reference to the functional evidence is also cited. Data on paralogue electrophysiology and disease associations is also included for triangulation (<https://scn-viewer.broadinstitute.org/>)^63^. Variant ID presented in vcf file format: *“chromosome: position: reference allele: alternative allele”.* Ephys GoF = Electrophysiology Gain-of-Function. EM = Erythromelalgia, PEPD = Paroxysmal Extreme Pain Disorder, SFN = Small Fibre Neuropathy, EP = Epilepsy. * = originally thought of as a possible EM mutation, but now considered to be a SFN mutation.

| Phenotype | Variable | Carriers | Carriers Pheno Rate | Controls | Controls Pheno Rate | Coef | SE | z | P | OR | L95CI | U95CI |
| --- | --- | --- | --- | --- | --- | --- | --- | --- | --- | --- | --- | --- |
| Chronic Pain | (Intercept) |  |  |  |  | 0.606 | 0.022 | -27.770 | 0.000 | 0.546 | 0.523 | 0.570 |
| Chronic Pain | Rare Carriers | 58 | 0.414 | 462158 | 0.418 | 0.034 | 0.267 | 0.127 | 0.899 | 1.035 | 0.613 | 1.746 |
| Chronic Pain | age |  |  |  |  | 0.006 | 0.000 | 17.029 | 0.000 | 1.006 | 1.006 | 1.007 |
| Chronic Pain | sex |  |  |  |  | -0.207 | 0.006 | -33.932 | 0.000 | 0.813 | 0.803 | 0.823 |
| Chronic Pain | PC1 |  |  |  |  | 0.001 | 0.000 | 10.390 | 0.000 | 1.001 | 1.000 | 1.001 |
| Chronic Pain | PC2 |  |  |  |  | 0.000 | 0.000 | -0.529 | 0.597 | 1.000 | 1.000 | 1.000 |
| Chronic Pain | PC3 |  |  |  |  | 0.003 | 0.000 | 12.615 | 0.000 | 1.003 | 1.002 | 1.003 |
| Chronic Pain | PC4 |  |  |  |  | 0.001 | 0.000 | 3.396 | 0.001 | 1.001 | 1.000 | 1.002 |
| Chronic Pain | PC5 |  |  |  |  | 0.002 | 0.000 | 5.283 | 0.000 | 1.002 | 1.001 | 1.003 |
| Chronic Pain | PC6 |  |  |  |  | 0.003 | 0.001 | 3.819 | 0.000 | 1.003 | 1.001 | 1.004 |
| Chronic Pain | PC7 |  |  |  |  | 0.004 | 0.001 | 5.960 | 0.000 | 1.004 | 1.002 | 1.005 |
| Chronic Pain | PC8 |  |  |  |  | 0.003 | 0.001 | 5.172 | 0.000 | 1.003 | 1.002 | 1.005 |
| Chronic Pain | PC9 |  |  |  |  | 0.001 | 0.001 | 1.809 | 0.070 | 1.001 | 1.000 | 1.003 |
| Chronic Pain | PC10 |  |  |  |  | 0.003 | 0.001 | 4.520 | 0.000 | 1.003 | 1.002 | 1.005 |

**Supplementary Table 6** – Logistic regression model summary for the effect of carrying a rare Na_V_1.7 mutation (Q10R, P187L, N641Y, R996C, M1532I) on the risk of chronic pain. *P* values presented in this table are unadjusted for multiple comparisons.

| Phenotype | Variable | Carriers | Carriers Pheno Rate | Controls | Controls Pheno Rate | Coef | SE | z | P | OR | L95CI | U95CI |
| --- | --- | --- | --- | --- | --- | --- | --- | --- | --- | --- | --- | --- |
| Chronic Pain | (Intercept) |  |  |  |  | -0.596 | 0.023 | -25.856 | 0.000 | 0.551 | 0.527 | 0.577 |
| Chronic Pain | All Carriers | 40236 | 0.418 | 462158 | 0.418 | -0.005 | 0.011 | -0.495 | 0.621 | 0.995 | 0.974 | 1.016 |
| Chronic Pain | age |  |  |  |  | 0.006 | 0.000 | 17.524 | 0.000 | 1.006 | 1.006 | 1.007 |
| Chronic Pain | sex |  |  |  |  | -0.205 | 0.006 | -35.091 | 0.000 | 0.814 | 0.805 | 0.824 |
| Chronic Pain | PC1 |  |  |  |  | 0.001 | 0.000 | 10.261 | 0.000 | 1.001 | 1.000 | 1.001 |
| Chronic Pain | PC2 |  |  |  |  | 0.000 | 0.000 | -0.945 | 0.345 | 1.000 | 1.000 | 1.000 |
| Chronic Pain | PC3 |  |  |  |  | 0.003 | 0.000 | 13.289 | 0.000 | 1.003 | 1.002 | 1.003 |
| Chronic Pain | PC4 |  |  |  |  | 0.001 | 0.000 | 3.816 | 0.000 | 1.001 | 1.001 | 1.002 |
| Chronic Pain | PC5 |  |  |  |  | 0.002 | 0.000 | 5.748 | 0.000 | 1.002 | 1.001 | 1.003 |
| Chronic Pain | PC6 |  |  |  |  | 0.002 | 0.001 | 4.002 | 0.000 | 1.002 | 1.001 | 1.003 |
| Chronic Pain | PC7 |  |  |  |  | 0.004 | 0.001 | 6.001 | 0.000 | 1.004 | 1.002 | 1.005 |
| Chronic Pain | PC8 |  |  |  |  | 0.004 | 0.001 | 6.091 | 0.000 | 1.004 | 1.002 | 1.005 |
| Chronic Pain | PC9 |  |  |  |  | 0.001 | 0.001 | 1.943 | 0.052 | 1.001 | 1.000 | 1.003 |
| Chronic Pain | PC10 |  |  |  |  | 0.003 | 0.001 | 4.288 | 0.000 | 1.003 | 1.002 | 1.004 |

**Supplementary Table 7** – Logistic regression model summary for the effect of carrying any proposed pathogenic Na_V_1.7 mutation (or combination thereof) on the risk of chronic pain. *P* values presented in this table are unadjusted for multiple comparisons
